# Improved Sleep, Cognitive Processing and Enhanced Learning and Memory Task Accuracy with Yoga Nidra Practice in Novices

**DOI:** 10.1101/2023.01.27.23284927

**Authors:** Karuna Datta, Anna Bhutambare, VL Mamatha, Yogita Ankush Narawade, Srinath Rajagopal, Madhuri Kanitkar

## Abstract

**Background:** Yoga nidra practice in novices is known to improve sleep. Its effect on objective parameters on sleep and on cognitive performance is not well known. The aim of the study was to study the effect of yoga nidra practice on cognition and night time sleep using objective parameters.

**Methods:** 41 healthy volunteers were enrolled and baseline sleep diary collected. Subjects underwent overnight polysomnography and cognition testing battery comprising of Motor praxis test (MPT), emotion recognition task (ERT), digital symbol substitution task (DSST), visual object learning task (VOLT), abstract matching (AIM), line orientation task (LOT), matrix reasoning task (MRT), fractal-2-back test (NBACK), psychomotor vigilance task (PVT-10 min) and balloon analog risk task (BART). Yoga nidra was practiced for two weeks after training. Cognition testing battery was done at baseline and at one and two weeks of practice to compare. The cognitive tasks were further analysed using Python library and power spectra density values (PSD) calculated for EEG frequencies at central, frontal and occipital locations. Repeat sleep diary and polysomnography to assess pre-post yoga nidra intervention effects were studied.

**Results:** Improved reaction times for all 10 cognition tasks was seen. Polysomnography (PSG) revealed significant difference in post intervention as compared to baseline. Data in change (95%CI; p-value) showed change in sleep efficiency, wake after sleep onset and delta μV^2^ in deep sleep : +3.62% (0.3, 5.15; p-value=0.03), -20min (−35.78, -5.02; p=0.003) and +4.19 (0.5, 9.5; p=0.04) respectively. Accuracy was found to be significantly increased for VOLT (95% CI: 0.08, 0.17; p=0.002), AIM (95% CI: 0.03, 0.12; p= 0.02) after two weeks of practice and NBACK (95% CI: 0.02, 0.13; p=0.04) with one week of yoga nidra practice. ERT accuracy scores with yoga nidra practice showed increased recognition scores in happy, fear and anger stimuli (95% CI: 0.07, 0.24; p=0.004) but reduced scores with neutral stimuli (95% CI: -0.3, -0.05; p=0.04) after two weeks of yoga nidra practice.

**Conclusion:** Yoga nidra practice improves cognitive processing and helps improve night-time sleep in healthy novices.

## Introduction

Yoga nidra practice has been associated with stress reduction, and has been used for insomnia, and other diseases ^1,2^. Good sleep quality is essential for human performance. Individuals required to take critical decisions require optimized cognitive functioning. There is evidence that improved sleep can enhance cognitive functioning and partial sleep deprivation reduces it ^3^.

Yoga nidra is documented in ancient scriptures as awake aware sleep. Methodology for its practice by novices is already laid out ^4^. Yoga nidra is proven to improve sleep in insomnia patients^1,5^. Subjective improvement of night-time sleep in healthy novices is also reported with evidence of local sleep during the practice in the morning. However its effect on objective parameters on sleep in healthy novices is not documented. Also whether yoga nidra practice can improve cognitive performance is also not documented.

There was a felt need to study the effect of yoga nidra practice in novices and study its effect on sleep and cognition objectively.

## Methods

### Study Design and Participants

The project was a pre-post interventional study with an aim to study the effect of yoga nidra practice on cognitive function and sleep. The study was conducted on healthy volunteers and their cognition was assessed at baseline and after yoga nidra practice. Objective sleep parameters using overnight polysomnography were also studied at baseline and after yoga nidra practice.

Yoga nidra supervised training model for novices developed by Datta et al in 2017,^5^ was used to study the effects on cognitive function and on night sleep. Participants were assessed after one and two weeks of practicing yoga nidra. Details of study design are given in Figure 1(a) and 1(b) for both objectives, i.e. the effect of yoga nidra practice on cognitive function and objective night-time sleep parameters respectively.

**Figure 1:**
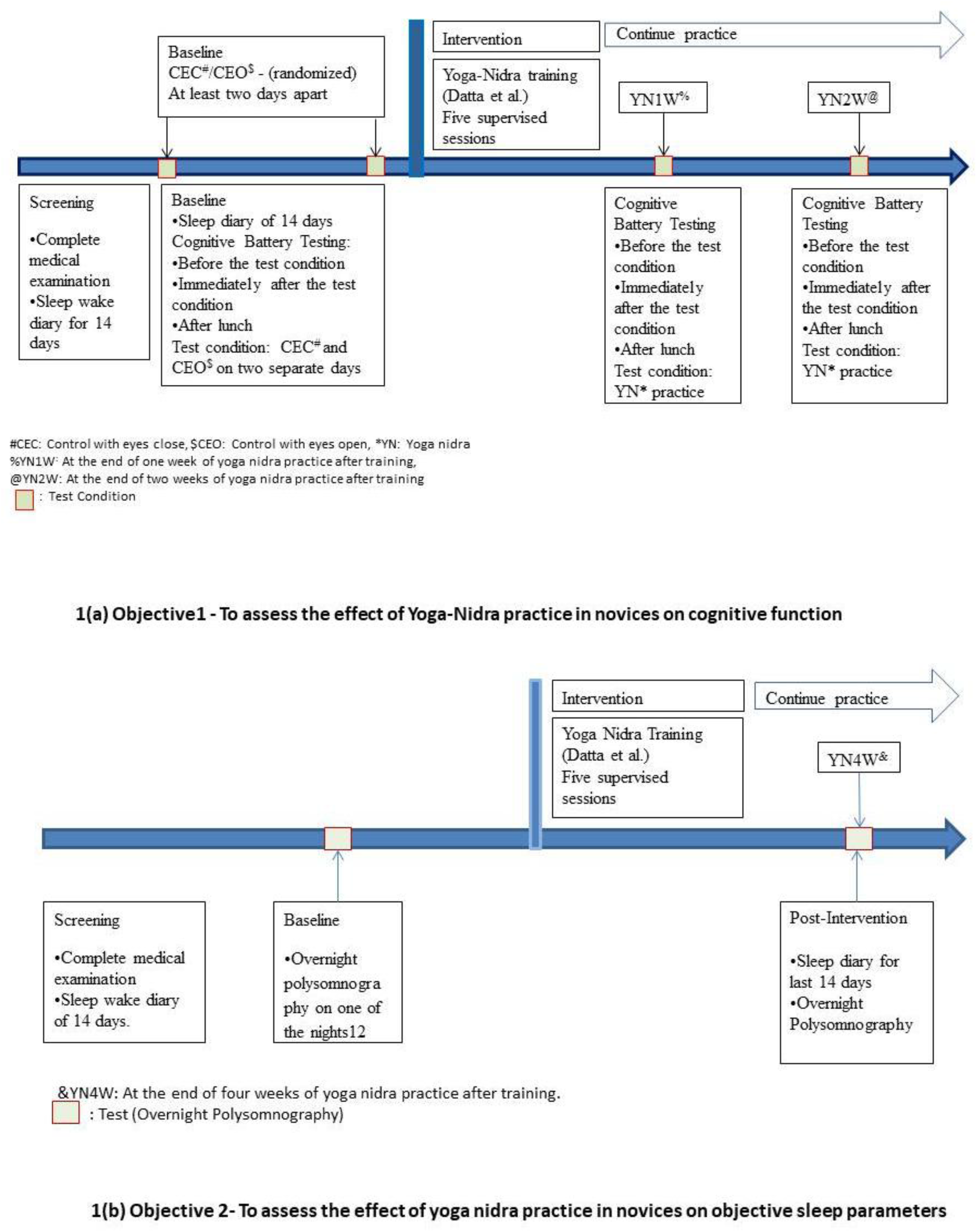
Study Design for effect of yoga nidra practice on cognitive function (Figure 1a) and on sleep (Figure 1b)

The study was conducted during 2021-2022 at a premier institute of India after institutional ethical clearance (IEC/2019/255 dated 25 Apr 2019). The study was registered in CTRI.in, vide no. CTRI/2021/031349. Local advertisements in the city were placed and volunteers assessed for eligibility. A minimum of 30 subjects for each objective was considered for this pilot study.

### Screening of Subjects & Enrollment

Healthy young male volunteers with a consistent sleep wake schedule were recruited. All subjects underwent a complete thorough examination by a medical practitioner. Sleep wake schedule was monitored and a baseline sleep diary of 14 days obtained. Exclusion criteria consisted of any sleep disorder, depression, h/o psychiatric disorder, and h/o acute illness which was likely to affect sleep wake schedule.

### Yoga-Nidra Session

A pre-recorded yoga nidra audio was used (*available at https://youtu.be/0fmMlELn-Ug*). Each Yoga-Nidra session consisted of seven steps, namely: preparation, *samkalpa* (resolution), body part awareness, breath awareness, feeling and sensation, visualization, and ending of practice. Supervised Yoga-Nidra sessions were done as per the therapeutic model developed earlier ^5^. During these supervised sessions, the investigator followed the ‘guidelines for observer’ developed for practice^1^.

### Methodology for assessing the effect of yoga nidra practice in novices on cognitive function

#### Cognitive Test Battery (CTB)

The cognition test battery was done using the Joggle Research Platform (USA). Our paradigm consisted of ten tests in a standard sequence of Motor Praxis Task(MPT), Visual Object Learning Task(VOLT), Fractal-2-Back(NBACK), Abstract Matching(AIM), Line Orientation Task(LOT), Emotion Recognition Task(ERT), Matrix Reasoning Task(MRT), Digital Symbol Substitute Task(DSST), Balloon Analog Risk Task(BART), and Psychomotor Vigilance Task(PVT:10min). The various cognition domains that were assessed by these 10 tests were: MPT – Sensory motor speed, VOLT – Spatial learning and memory, NBACK – Working memory, AIM – Abstraction and concept information, LOT – Spatial orientation, ERT – Emotion identification, MRT – Abstract reasoning, DSST – Complex scanning and visual tracking, BART – Risk decision making, and PVT – Vigilant attention^6^.

CTB was performed on the test condition days of control with eyes open (CEO), control with eyes close (CEC), at the end of 1^st^ week of Yoga nidra practice (YN1W), and at the end of 2^nd^ week of yoga nidra practice (YN2W). All the subjects completed the CTB paradigm tests three times on each test condition day i.e. at 10 ‘o’ clock (Mr1), immediately after the test condition of CEC, CEO, YN1W, or YN2W (Mr2) and then in the afternoon at 1500h (post lunch: PL). The conditions were kept standardized by similar food, working environment on all condition days for each subject. Detailed representation of study design is shown in Figure 1(a).

CTB outcome parameters studied were reaction time (for all 10 tests) and accuracy score for 9 tests except BART. For BART –’Adjusted Number of Pumps’, ‘Number of Explosions’, ‘Risk Propensity’ and ‘Cash collected’ were analyzed^7^. ‘Adjusted Pumps’ is the average number of pumps on each balloon during BART when the balloon did not burst, ‘Number of Explosions’ is the total number of balloons exploded during BART, ‘Risk Propensity’ is the ratio of total pumps/balloons collected, and ‘Cash collected’ is the amount of reward earned in BART ^6^. The standard accuracy outcome was ‘proportion correct’ ranging from ‘zero’ to ‘one’, where ‘zero’ and ‘one’ represented worst and best possible performance respectively. For the MPT, the distance from the center of the square to the point where participant touches is used as an accuracy score. The center of square translates to ‘one’ accuracy score and an edge of square translate to ‘zero’ accuracy score, with linear scaling between center and edges. For LOT, the accuracy score was calculated as ‘three’ minus the average number of clicks off, which was then divided by ‘three’. For tests with average number of clicks off is more than ‘three’, the accuracy score was set to ‘zero’. For PVT, the accuracy score was calculated as ‘one’ – [(number of lapses + number of false starts) / (number of stimuli + number of false starts)]. PVT-number of false starts was analyzed apart from reaction time and accuracy^6^. ERT was also studied for various types of stimuli apart from reaction time and accuracy.

#### Data Analysis

One-Way Repeated Measures (RM) Analysis of Variance (ANOVA) for checking significant change in each of the outcome variable between different test conditions (CEO, CEC, YN1W, and YN2W) and at different times (Mr1, Mr2, or PL) used for normal data. If data was not normal, Friedman’s test was used. In instances where the sphericity assumption was not met, the reported p-values associated with the F-statistics were adjusted via the Greenhouse-Geisser correction. If the ANOVA model was significant, post-hoc analysis (pairwise comparison) was done using paired t-test with Bonferroni Correction. Wilcoxon Signed Rank Test with Bonferroni Correction was used for post-hoc analysis (pairwise comparison) if Friedman’s Test was significant. Bonferroni Correction was used to reduce the probability of type-1 error (i.e. chance of obtaining false-positive results). All reported p-values were two-tailed and statistical significance was assumed if p-value < 0.05. All Statistical analysis was done using R-software (version 4.1.2).

#### PSD Data of EEG daytime recording during CTB

Power Spectra Density (PSD) using continuous EEG data was obtained and PSD calculated from continuous daytime EEG data in Python. The spectral analysis of EEG data was performed using MNE-Python, an open-source library for visualizing, analyzing and exploring the raw EEG signal^8^. This included loading data, pre-processing, and segmentation of the continual data on the obtained epoched data. Time-Frequency investigation was performed and finally spectral-parameters of the EEG signal was extracted ^9^. Spectral investigation is mainly based on frequency bands. The Power Spectral Density (PSD) is the reflection of frequency components of EEG signal analyzing the patterns of the frequency bands^10^. The calculation of Power-Spectrum was carried out for the EEG data while doing the CTB for 12 participants. Preprocessing included low and high frequency filtering of the EEG signal, Notch filtering, removal of bad segments of the data (made by visual screening of data for sudden artifacts). Eye movements were repaired using the artifacts repair regression technique, custom Re-referencing and segmentation of the EEG data to epochs. Filtering was done using Low-Frequency 1Hz and the High-Frequency 90Hz. Notch-filter at 50Hz was used. After manual marking of artifacts and bad segments, MNE-Python library was used to exclude automatically annotated spans of data while creating the epochs from the continual EEG data. Special care was taken to avoid segments with artifacts or noise so that artifact free data would be extracted for further analysis. The EEG signal acquisition was carried based on the domain of time and future extraction was done. Slow eye movement artifacts can be recorded as EEG activity at the time of EEG signal acquisition and regression coefficients on obtained epoched EEG raw signal through subtracting the evoked response was taken. The computed coefficients were used to repair the artifacts and elimination of EOG signal done from EEG signal^11^.

Segmentation was performed on the clean raw EEG data to segment the EEG data to epochs. The Feature extraction was done based on Time-Frequency domain ^9, 12^. The time-frequency domain depicts the distribution of power of the EEG signal with respect to Time-Frequency plane. The computation of Power Spectral Density (PSD) was carried out using Welch’s Technique. The decomposition of signal to frequency bands were done using Fourier transforms ^9^. The ranges of the bands were delta (1-4Hz), theta1 (4-6Hz), theta2 (6-8Hz), alpha1 (8-10Hz), alpha2 (10-12.5Hz), beta (13-30Hz) ^5^. The features used in the EEG signal analysis were Relative-band-power and ratios between the bands ^13^. On each test segment spectral analysis was performed for all the EEG channels. Using 512ms window duration and 256Hz sampling rate the features were extracted on the desired test segments for all the EEG channels on every individual subjects. The specified spectral analysis methodology was employed for all the subjects for all test conditions i.e. CEC, CEO, YN1W, and YN2W for all the times i.e. Mr1, Mr2, and PL. PSD values were compared for various frequency bands and also as ratios. The various parameters thus studied are: delta, theta1, theta2, alpha1, alpha2, beta, delta/beta, theta1/beta, theta2/beta, theta1/alpha1, theta2/alpha1, theta1/alpha2, theta2/alpha2, delta/theta1, delta/theta2, delta/alpha1, and delta/alpha2.

##### Data Analysis

One-way ANOVA model was used for checking significance of difference in the PSD outcome parameters and post hoc analysis using Tukey’s post-hoc test if data was normal. Otherwise Kruskal Wallis test was used with Wilcoxon test for post hoc analysis. Statistical significance was considered at alpha 0.05. All Statistical analysis was done using R-software (version 4.1.2).

#### Sleep Diary (SD)

All subjects filled a baseline sleep diary for 14 days. The sleep diary was filled twice a day, once in the morning just after getting up and again at night just before bedtime. The subjects continued to fill the sleep diary during the intervention. The sleep diary parameters obtained were: time in bed (sdTIB), total sleep time (sdTST), sleep onset latency (sdSOL), wake after sleep onset (sdWASO), sleep quality (sdSQ), and sleep efficiency (sdSE)^5^. These parameters were extracted from the two week data of baseline and post-intervention.

##### Data Analysis

Sleep diary data was collected and analyzed for pre and post comparison. Normality of data was checked using Shapiro-Wilk’s test. Paired t-test was used to compare sleep diary parameters before (baseline) yoga nidra practice and after (post-intervention) yoga nidra practice, if data was normal. For non-normal data several transformations like: square, square root, log, exponential were tried. If normality was not achieved, Wilcoxon signed rank test was used. Statistical significance was considered at alpha 0.05. All Statistical analysis was done using R-software (version 4.1.2).

### Methodology for assessing the effect of yoga nidra practice in novices on objective sleep parameters

#### PSG for Objective Sleep Parameters

To assess the effect of yoga nidra on objective sleep parameter it was planned to do the overnight polysomnography (PSG) of 30 subjects was done. PSG was conducted according to AASM criteria^14^ using ‘SOMNOMEDICS©, Germany PSG System’. EEG, EOG, EMG channels were placed for scoring sleep-wake stages using DOMINO© software version 3.0.0.6. F3, F4, C3, C4, O1, and O2 EEG channels were placed as per the 10-20 system. PSG study was done during the night under standard conditions. The impedance was kept below 5KΩ. Polysomnography was performed as baseline (BL PSG) and post intervention i.e. four weeks of yoga nidra practice (PI PSG).

PSG data files were analyzed by KD in groups of 15-20 files and each was coded to blind KD from the identity state of subject data i.e. of baseline (BL) or post-intervention (PI). The files once analyzed were decoded for further analysis.

Various parameters obtained using PSG were: time in bed (TIB), total sleep time (TST), wake duration, wake after sleep onset (WASO), and duration of various stages of sleep (i.e. N1, N2, N3 and REM). EEG analysis i.e. alpha, beta, theta and delta waves amplitude in μV^2^. Percentages for sleep, wake, REM, and Non-REM sleep was calculated and also analyzed.

##### Data Analysis

Normality of data was checked using Shapiro-Wilk’s test for normality. Quantile -Quantile plot for each of the outcome parameter was made. If normality was not obtained even after square root, square, log, and exponential transformations then analysis using Wilcoxon Signed Rank Test was done. All statistical analysis was done using R-software (version 4.1.2).

## Results

Details of number of subjects screened, allocated, and finally analyzed is shown in Figure2. 12 subjects volunteered for daytime EEG recording during CTB. Sleep diary analysis is shown in Table 1. Polysomnography data (baseline; post-intervention; p-value) of TIB (min), TST (min), and Total Non-REM was (428·09±82·13; 423·02±72·96; 0·676), (372·36±68·46; 387·70±74·94; 0·241), and (340·73±111·95; 341·88±73·48; 0·953) respectively. Pre-post intervention analysis of other PSG parameters is shown in Figure 3. Cognition battery testing results for reaction time is shown in Figure 4(i) and (ii) and accuracy scores are shown in Figure 5(i) and (ii). Trend graphs for accuracy and reaction time for the CTB is given in Supplementary Figure 1 and Figure 2 respectively. BART results for ‘adjusted number of pumps’, ‘risk propensity’, ‘cash collected’, and ‘number of balloons burst’ is shown in Figure 6. PVT result of ‘number of false starts’ is shown in Figure 7. ERT analysis for various types of stimuli is shown in Supplementary Table 2. Significant test results in EEG analysis of power spectra density using Python are shown in Table 2.

**Table 1:**
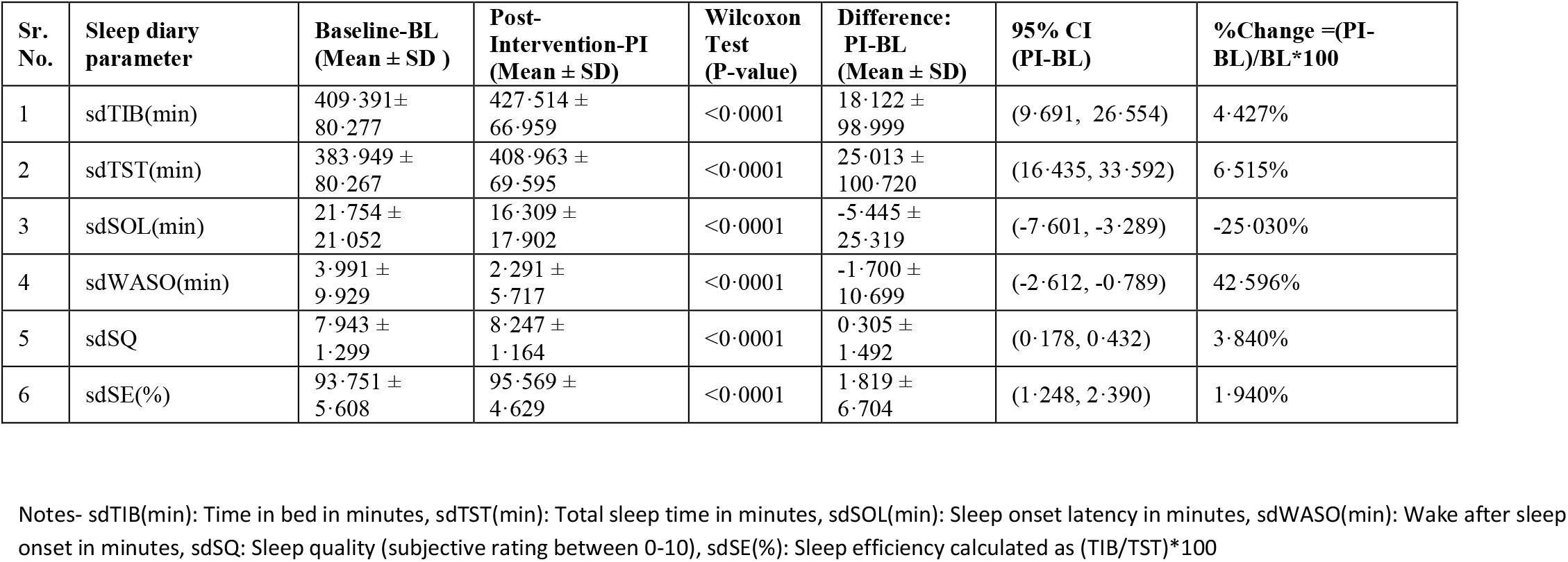
Sleep Diary Parameters of subjects before (baseline) and after yoga nidra practice (post intervention)

| Sr. No. | Sleep diary parameter | Baseline-BL (Mean ± SD ) | Post-Intervention-PI (Mean ± SD) | Wilcoxon Test (P-value) | Difference: PI-BL (Mean ± SD) | 95% CI (PI-BL) | %Change =(PI-BL)/BL*100 |
| --- | --- | --- | --- | --- | --- | --- | --- |
| 1 | sdTIB(min) | 409.391± 80.277 | 427.514 ± 66.959 | <0.0001 | 18.122 ± 98.999 | (9.691, 26.554) | 4.427% |
| 2 | sdTST(min) | 383.949 ± 80.267 | 408.963 ± 69.595 | <0.0001 | 25.013 ± 100.720 | (16.435, 33.592) | 6.515% |
| 3 | sdSOL(min) | 21.754 ± 21.052 | 16.309 ± 17.902 | <0.0001 | -5.445 ± 25.319 | (-7.601, -3.289) | -25.030% |
| 4 | sdWASO(min) | 3.991 ± 9.929 | 2.291 ± 5.717 | <0.0001 | -1.700 ± 10.699 | (-2.612, -0.789) | 42.596% |
| 5 | sdSQ | 7.943 ± 1.299 | 8.247 ± 1.164 | <0.0001 | 0.305 ± 1.492 | (0.178, 0.432) | 3.840% |
| 6 | sdSE(%) | 93.751 ± 5.608 | 95.569 ± 4.629 | <0.0001 | 1.819 ± 6.704 | (1.248, 2.390) | 1.940% |

Notes- sdTIB(min): Time in bed in minutes, sdTST(min): Total sleep time in minutes, sdSOL(min): Sleep onset latency in minutes, sdWASO(min): Wake after sleep onset in minutes, sdSQ: Sleep quality (subjective rating between 0-10), sdSE(%): Sleep efficiency calculated as (TIB/TST)\*100

**Table 2:**
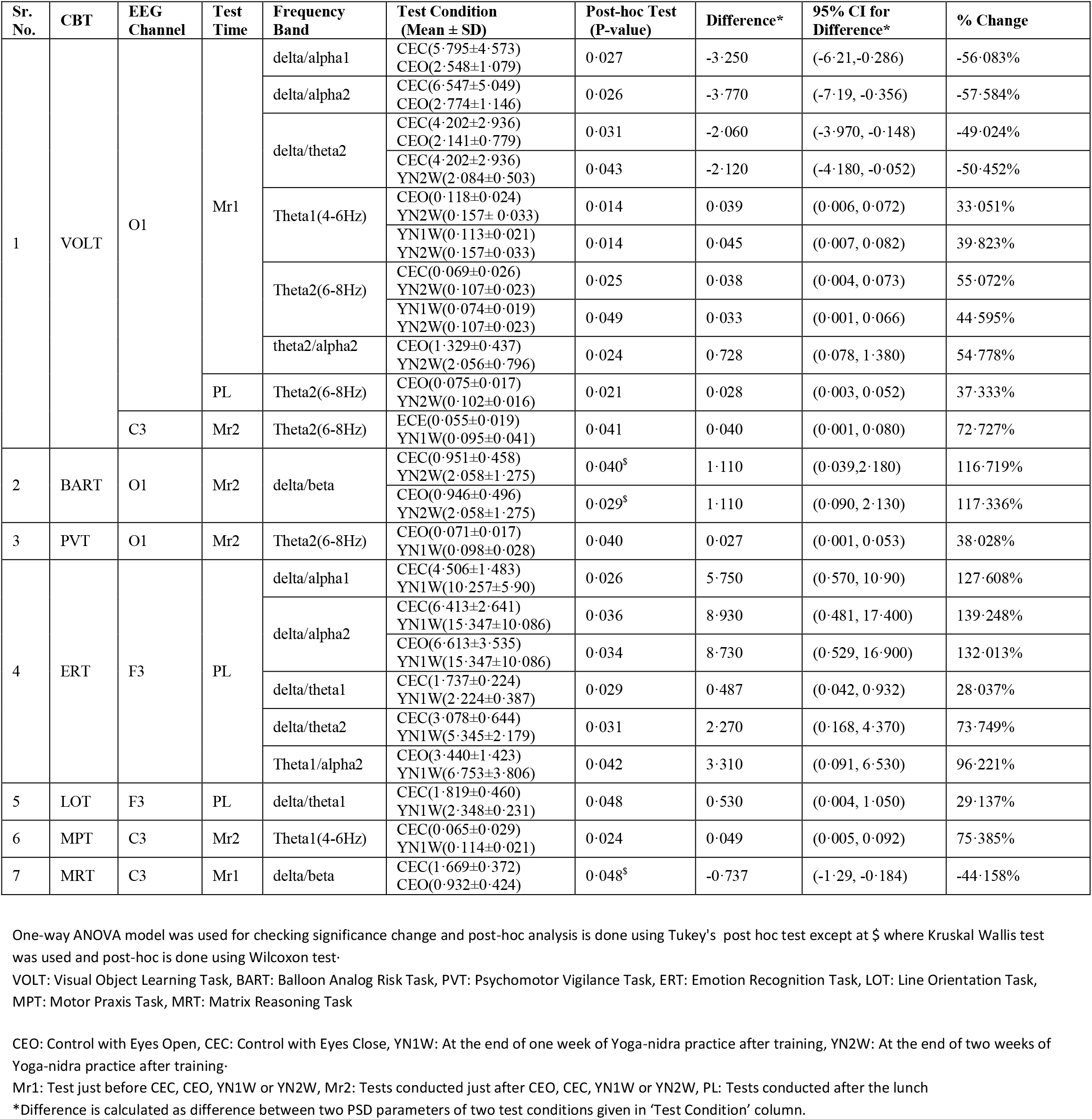
Significant daytime and test condition PSD results in frequency bands during CBT at various EEG locations.

**Figure 2:**
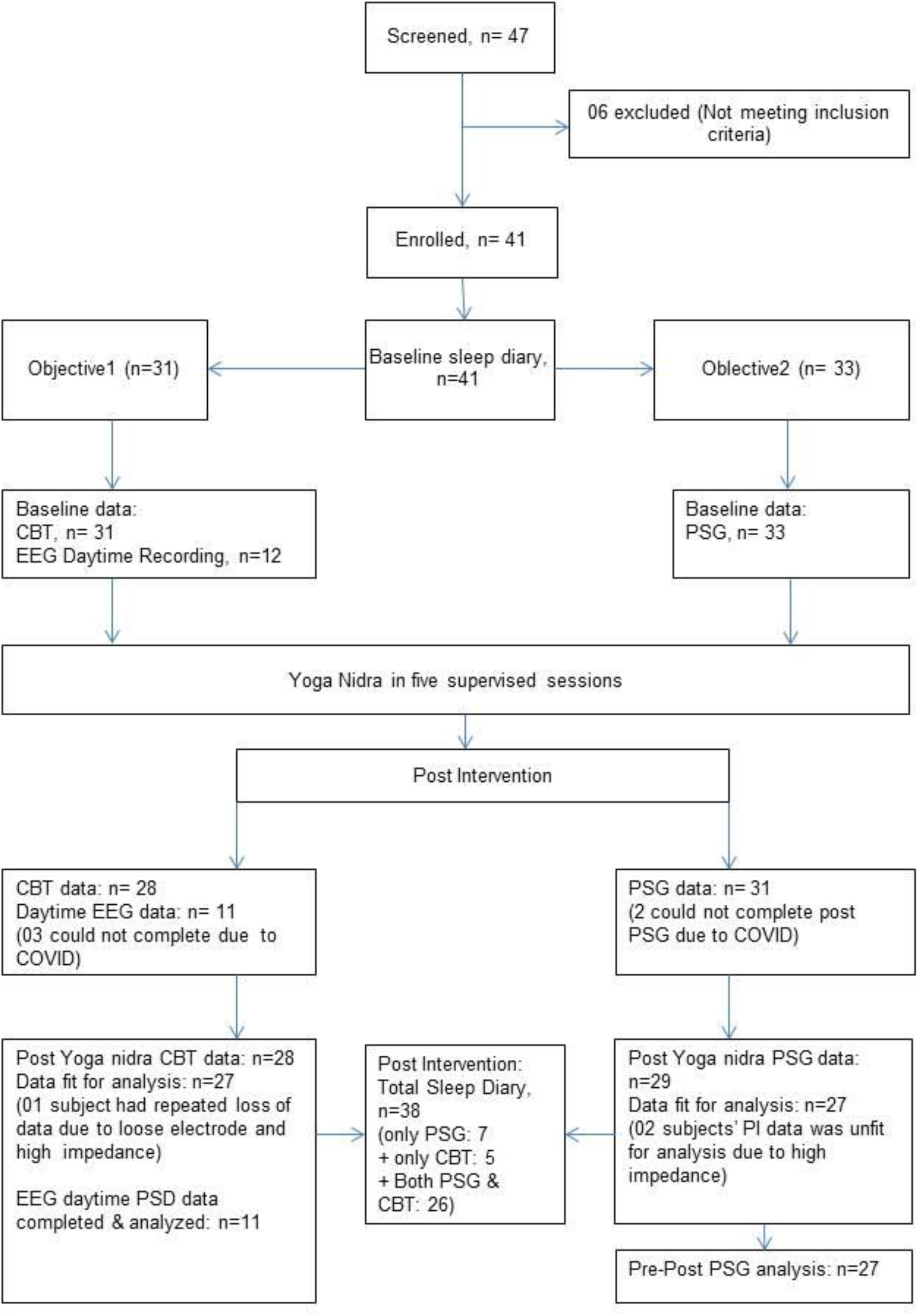
Details of the subjects screened, allotted and finally analyzed.

**Figure 3:**
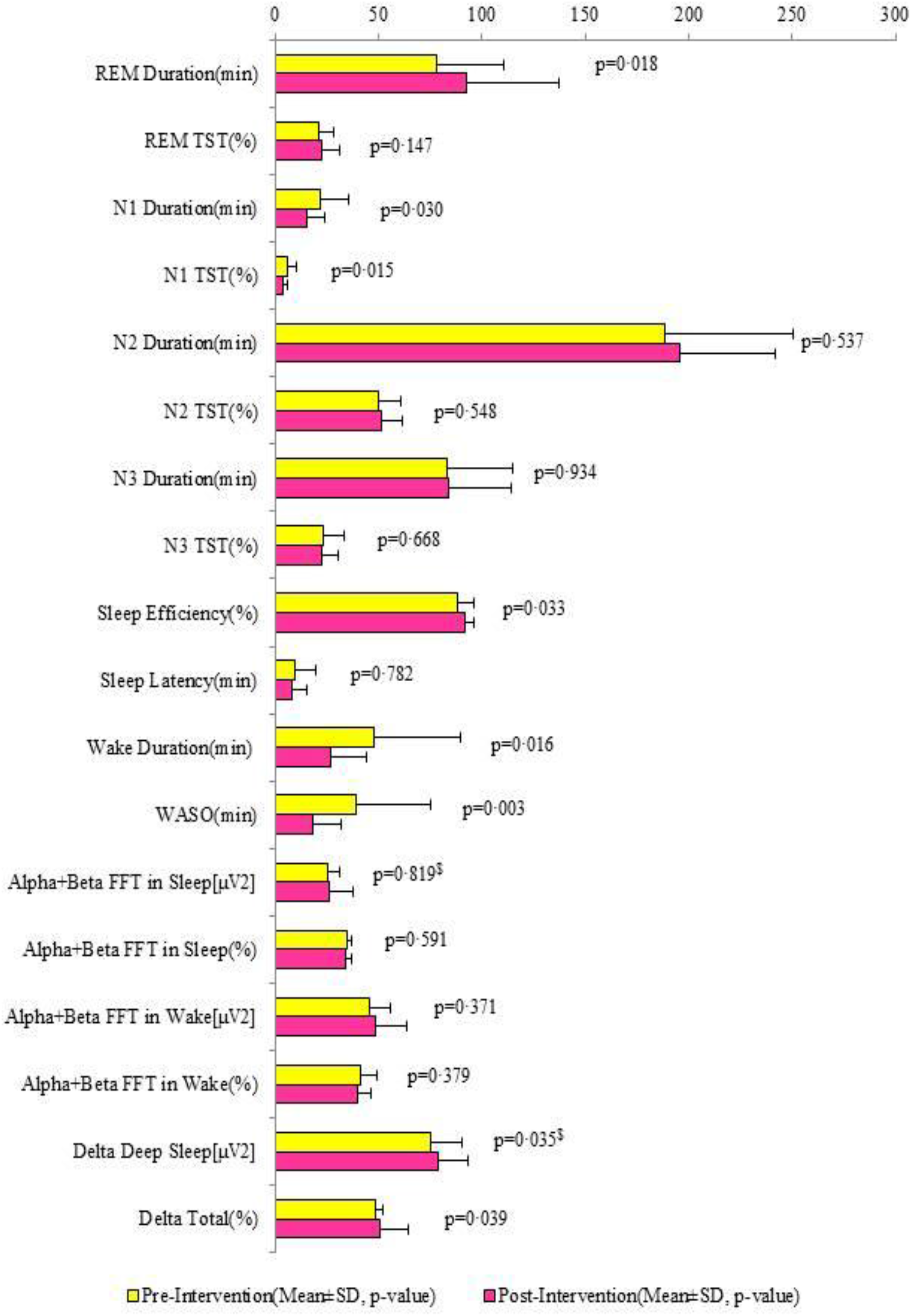
PSG Objective Sleep Parameters Comparison in Subjects before (baseline) and after (post-intervention) yoga nidra training. Note-$: p-values computed using Wilcoxon signed rank test. Rest all p-values are computed using two-tailed paired t test.

**Figure 4:**
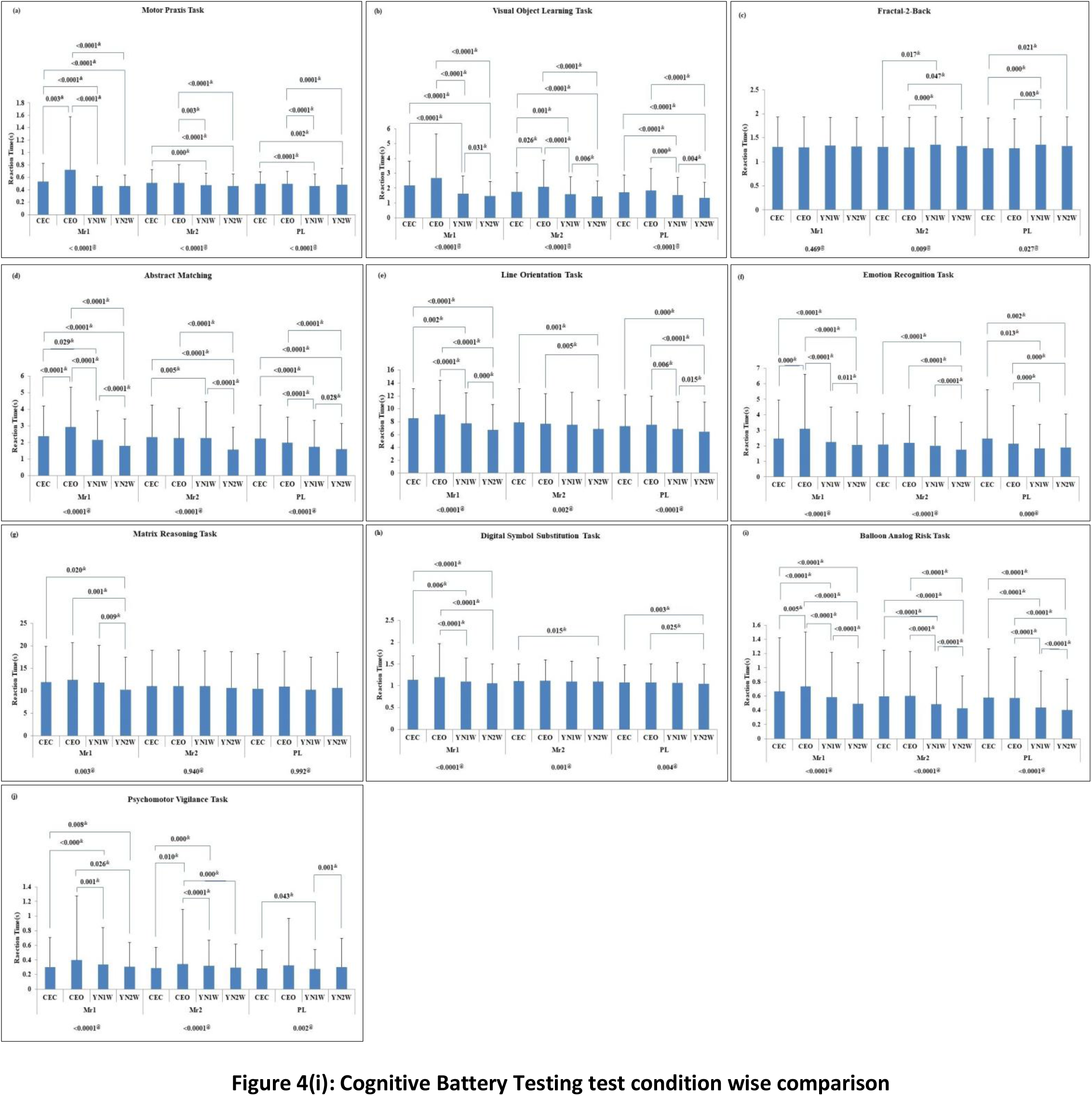
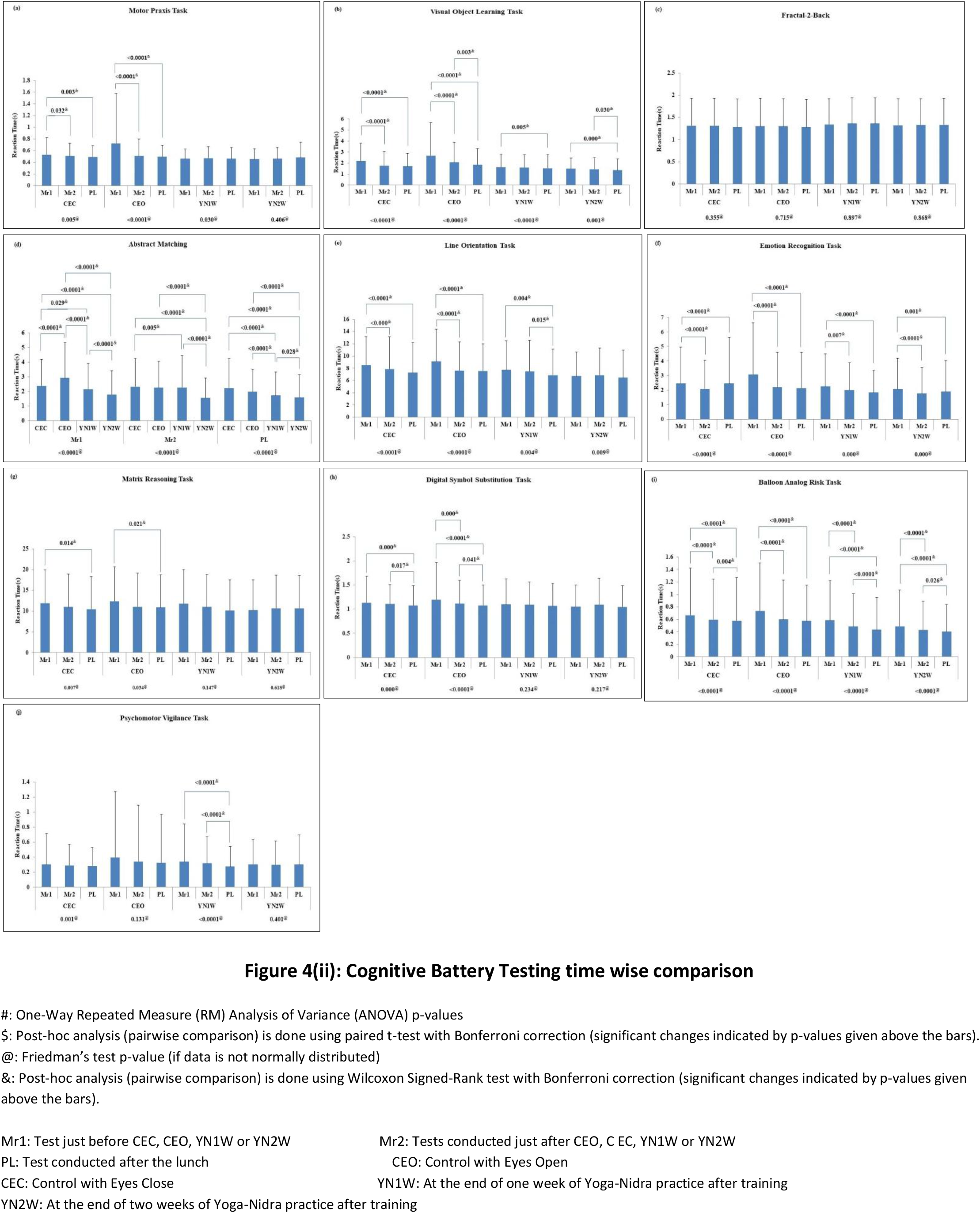
Cognitive Battery Test results showing reaction time (Mean ± SD) in (a)MPT, (b)VOLT, (c)NBACK, (d)AIM, (e)LOT, (f)ERT, (g)MRT, (h)DSST, (i)BART, and (j)PVT with test condition wise comparison (fig. 4(i)) and time wise comparison(fig. 4(ii)). (Comparison model for each test condition i.e. CEC, CEO, YN1W, and YN2W is shown below each time i.e. Mr1, Mr2, and PL. Significant post-hoc p-values are also in figure 4(i) and vice versa in 4(ii) depicted)

**Figure 5:**
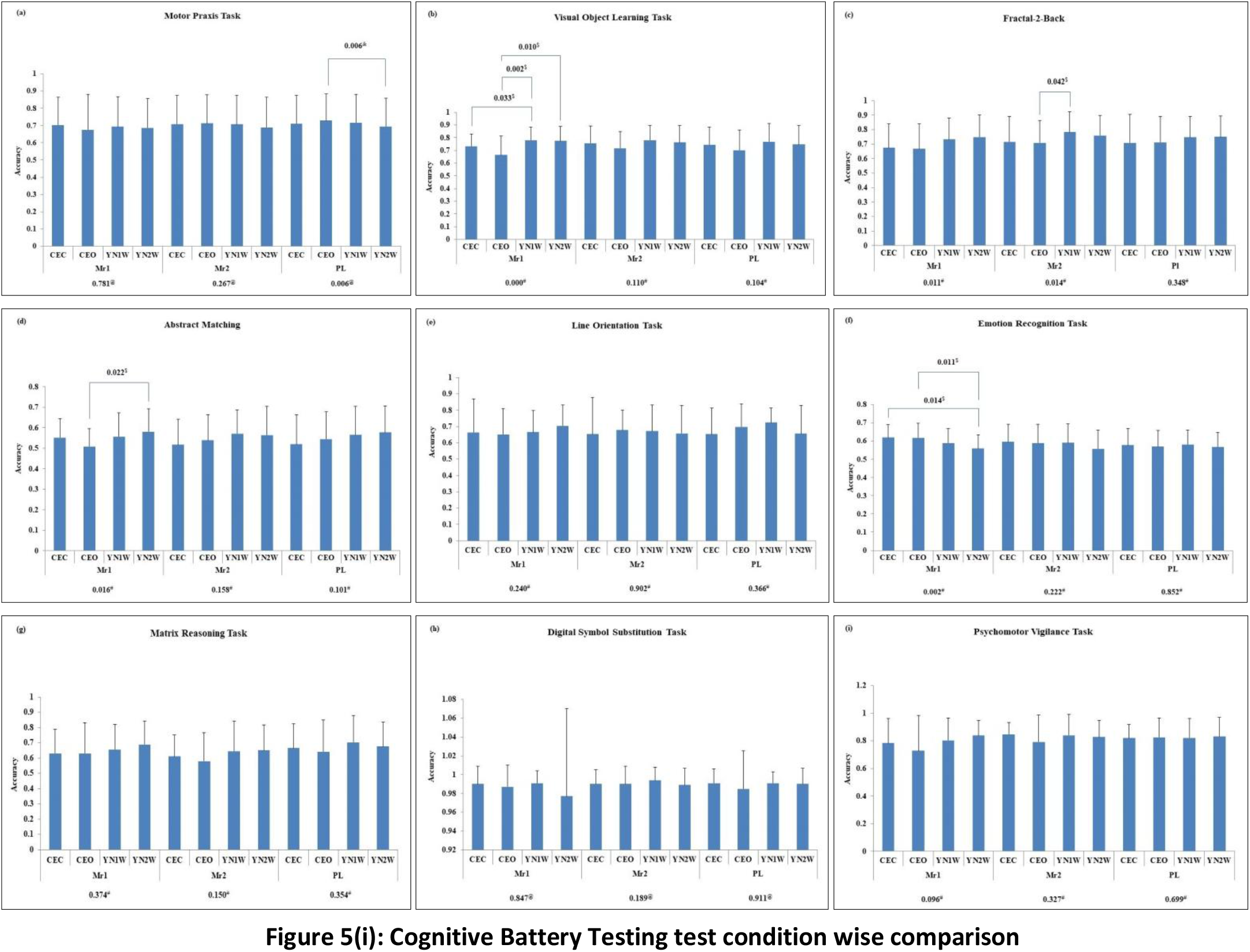
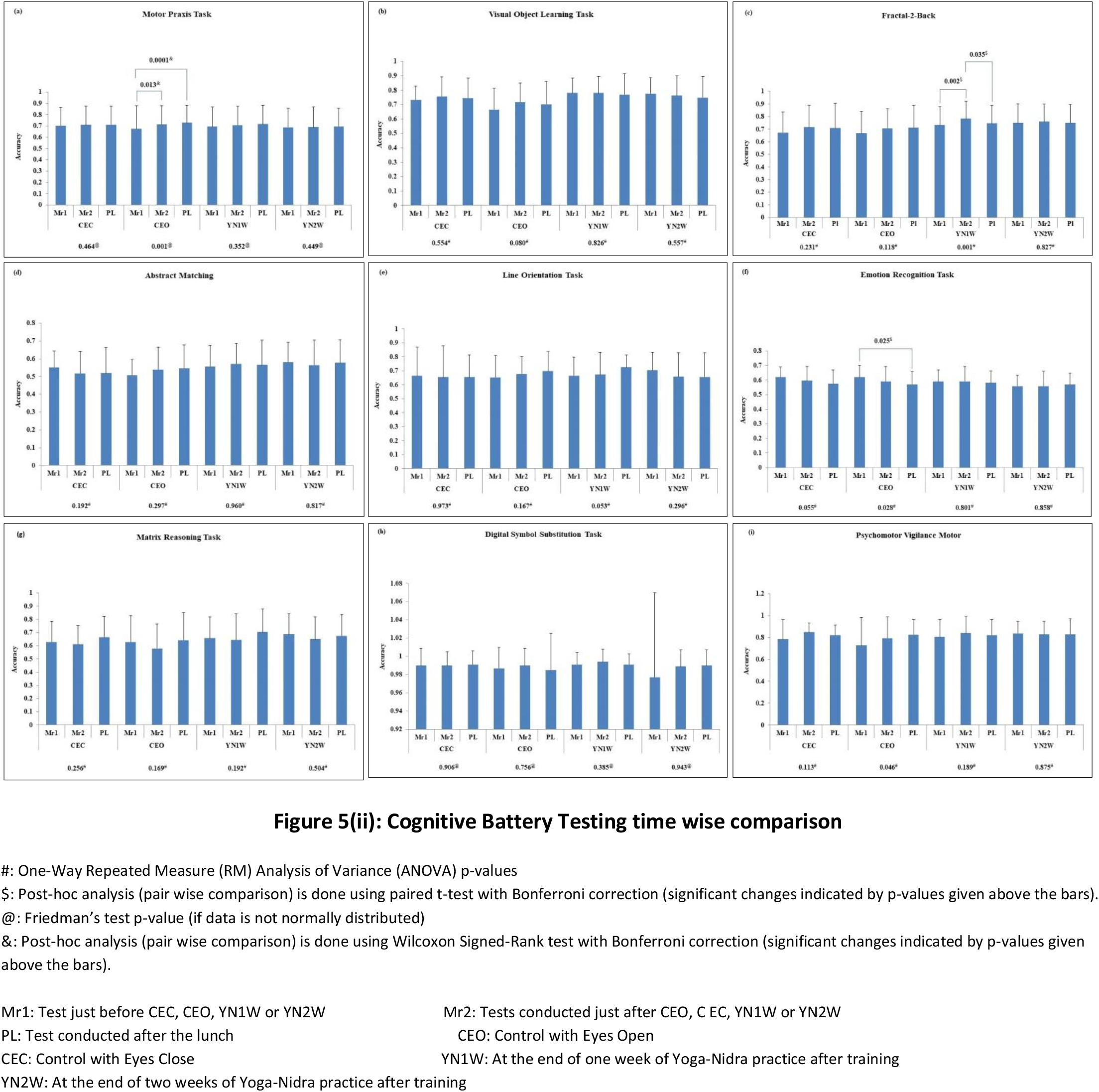
Cognitive Battery Test results showing accuracy measure (Mean ± SD) in (a)MPT, (b)VOLT, (c)NBACK, (d)AIM, (e)LOT, (f)ERT, (g)MRT, (h)DSST, and (i)PVT with time wise comparison(fig. 5(i)) and time wise comparison(fig. 5(ii)). (Comparison model for each test condition i.e. CEC, CEO, YN1W, and YN2W is shown below each time i.e. Mr1, Mr2, and PL. Significant post-hoc p-values are also in figure 4(i) and vice versa in 4(ii) are also depicted)

**Figure 6:**
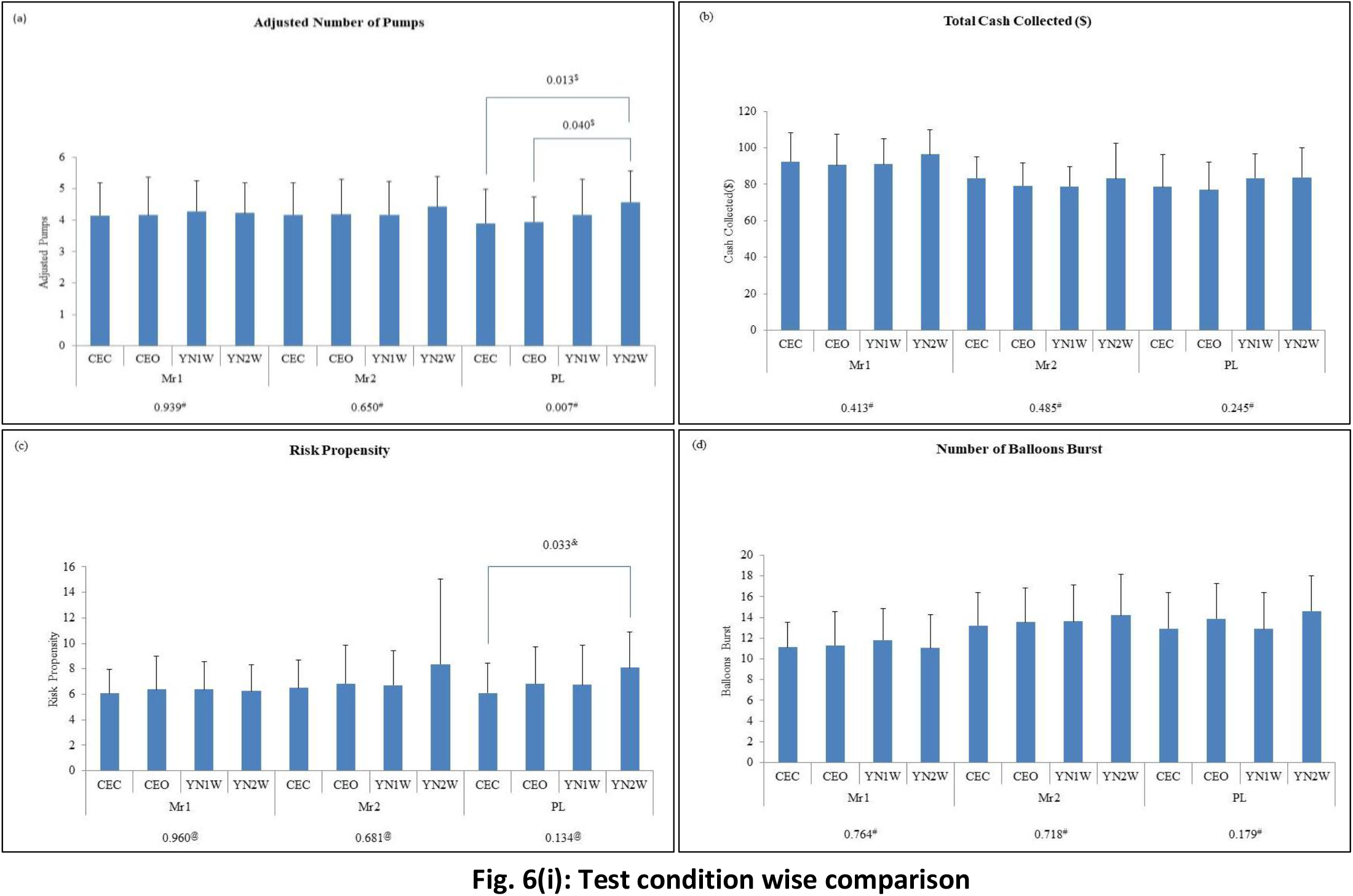
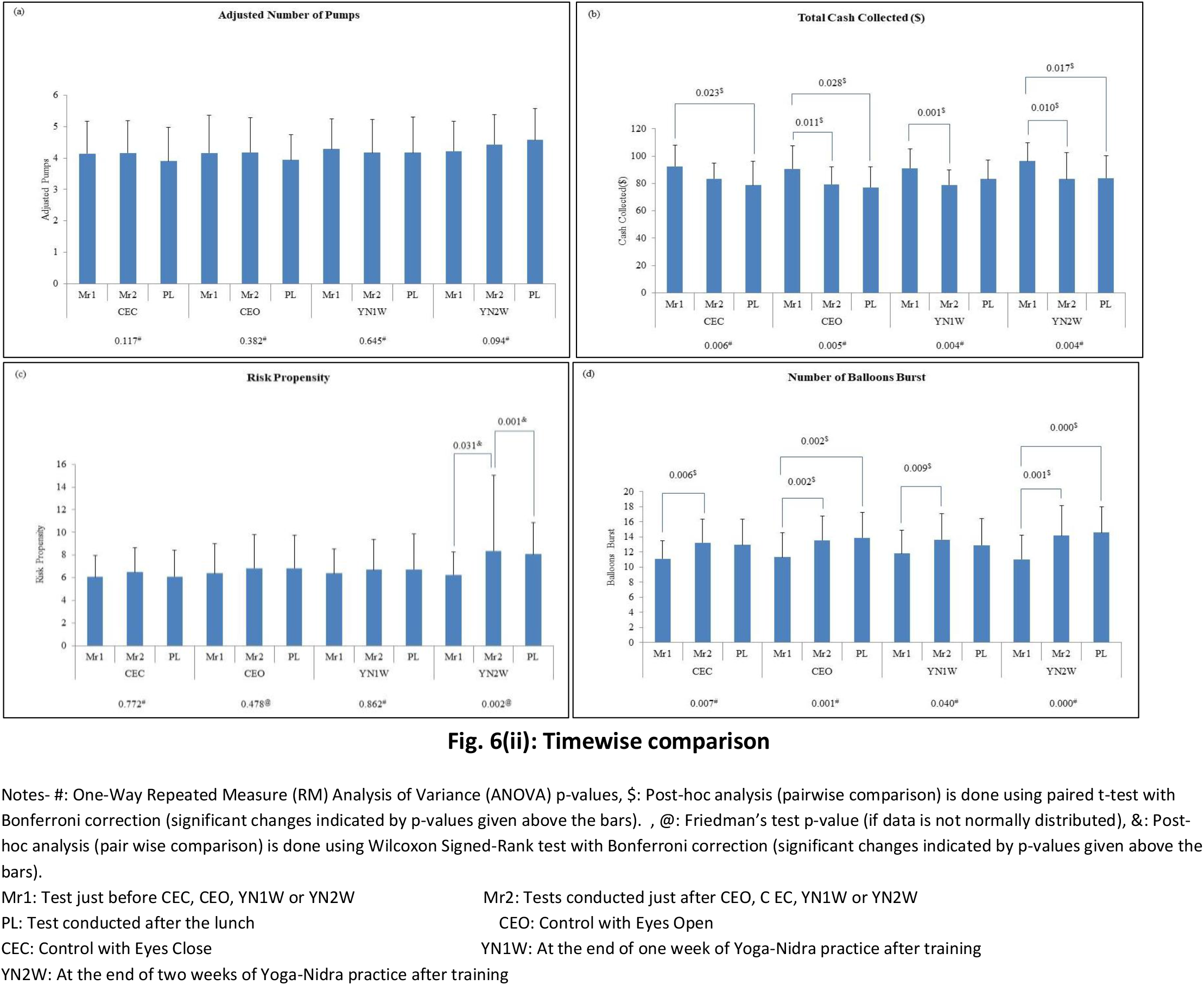
BART results (Mean ± SD) of a) Adjusted Number of Pumps, b)Total cash collected($), c) Risk Propensity, and d) Number of Balloons burst before (baseline) and after (post-intervention) yoga nidra practice test condition wise (Fig. 6(i)) and time wise(Fig. 6(ii)). (P-values for time wise and test condition wise comparison model is shown below i.e. for Mr1, Mr2, and PL; and CEC, CEO, YN1W, and YN2W respectively. Significant post-hoc p-values are also depicted)

**Figure 7:**
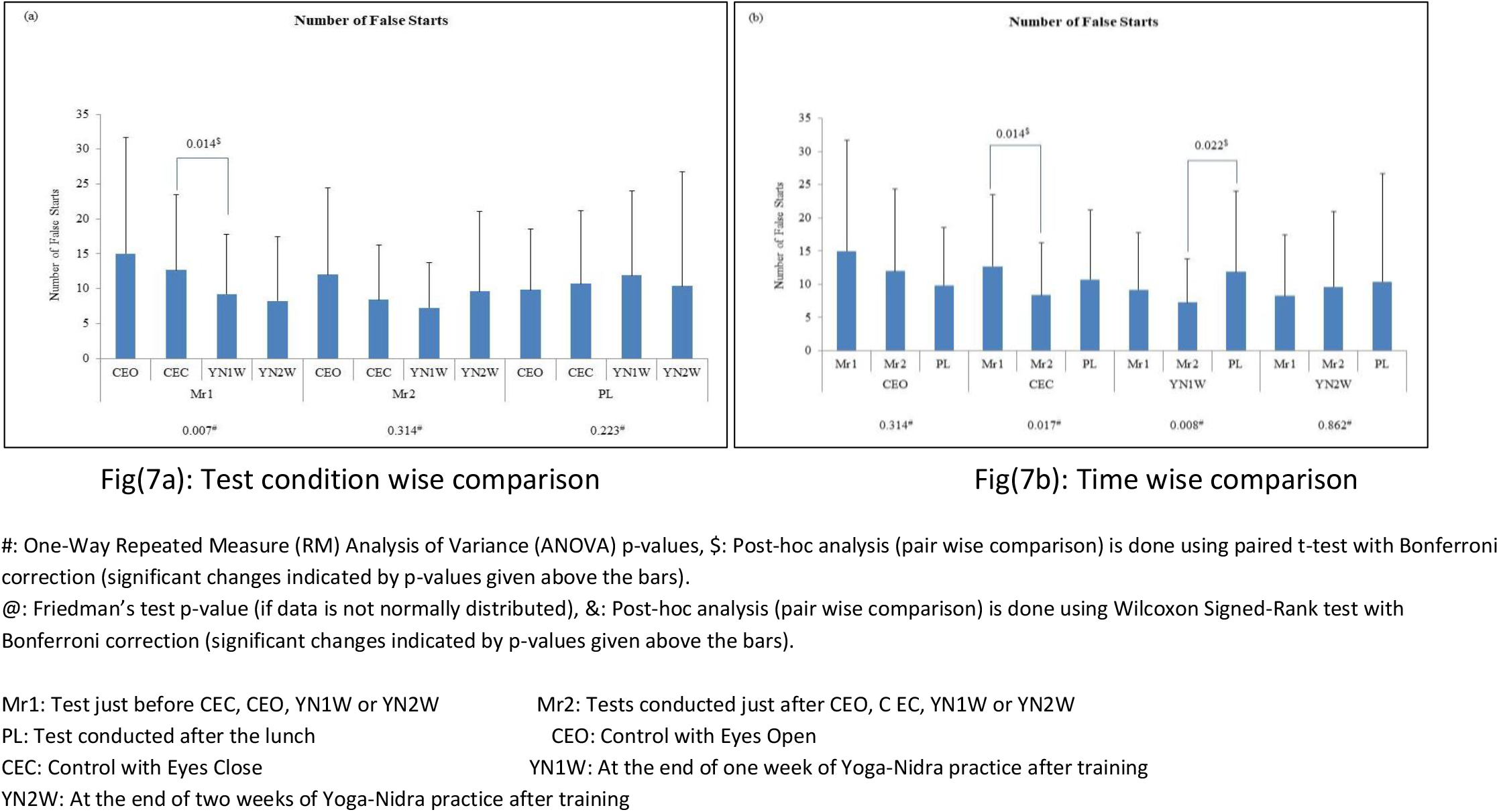
PVT-Number of False Starts (Mean ± SD) of subjects before (baseline) and after (post-intervention) yoga nidra practice comparison in test condition wise (Fig. 7a) and time wise (Fig. 7b) (Comparison model for each test condition i.e. CEC, CEO, YN1W, and YN2W is shown below each time i.e. Mr1, Mr2, and PL. Significant post-hoc p-values are also in figure 7a) and vice versa in 7b) are also depicted)

## Discussion

Yoga nidra practice by novices is known to improve sleep in insomnia patients ^1,2,5,15^. In healthy subjects the practice showed evidence of local sleep in certain areas of brain while the subject was behaviorally awake ^3^. Our results showed an objective improvement in night time sleep after yoga nidra practice in novices. The mechanism of improvement in night time sleep with yoga nidra done during the morning hours is probably due to the reduction in the sympathetic drive and increase in parasympathetic drive ^16^. However the exact mechanism is still not elucidated. We found a significant improvement in total sleep time (TST), sleep quality (SQ), sleep onset latency (SOL), and WASO which was reported subjectively by the patients using sleep diary. Objectively we found an increase in sleep efficiency, improvement in WASO and a significant improvement in delta sleep (%) of slow wave sleep. This objective improvement of the delta sleep in slow wave sleep is an important factor in improvement of sleep quality. Enhancement of slow wave sleep is reported using auditory stimuli during sleep, gaboxadol, tiagabine and transcranial direct cranial stimulation^17–19^. Slow wave sleep enhancement has been found to improve attention, learning, memory and performance^15,18,19^. Slow waves during sleep have been directly or indirectly linked to synaptic strength as a part of synaptic homeostasis theory ^20^. Local sleep seen during the yoga nidra practice is also found to enhance task performance ^3,21^. We found enhanced reaction time of all tests with no reduction in accuracy after the practice as compared to baseline. The local sleep seen in central, occipital and parietal areas during the rotation of consciousness in yoga nidra practice as brought out Datta et al earlier may be as a result of practice. Though not studied in this study, but probably increase in synaptic strength as per the synaptic hypothesis might be responsible^20^

Yoga nidra practice showed improvement in reaction time with no deterioration in accuracy of all cognitive battery tests i.e. MPT, VOLT, NBACK, AIM, LOT, ERT, MRT, DSST, BART, and PVT. This implies an increase in processing speed. Certain tests like AIM, ERT, NABCK, and VOLT actually showed an increase in accuracy too. Implying effects of yoga nidra practice directly or indirectly on the various brain regions involved in the tests.

To understand the mechanism of the effect of the yoga nidra practice during these cognitive tasks EEG signals at O1, F3, and C3 showed significant changes in VOLT, where the accuracy was also found to be increased. The delta and theta at O1 was more in YN2W also a significant increase at C3 immediately after yoga nidra practice, implying a local sleep type phenomenon at this area while improvement in accuracy implying a probable direct or indirect effect on middle-temporal cortex and hippocampus, areas associated with VOLT implying increased spatial learning in memory. Increased delta propensity during tasks as found using analysis on EEG PSD values, especially in the early morning readings after two weeks of yoga nidra practice showed a probable effect of a not so immediate effect of practice or rather a post night sleep effect after yoga nidra practice the previous few days. After the practice also delta propensity continued without reduction inaccuracy rather improving reaction time.

Accuracy improved in Abstract Matching test which is specifically related to prefrontal cortex and includes Abstraction and concept formation. This accuracy found to be increased early morning which may be as a direct or indirect effect related to improved sleep after yoga nidra practice. Slow wave sleep is essential for enhanced prefrontal cortex activity. Working memory as shown with NBACK test also showed significant improvement with yoga nidra practice. The actual number of ‘adjusted number of pumps’ which is on those balloons which were collected showed a significant increase in post lunch session after two weeks of practice.

ERT showed a significant deterioration in the early morning task scores after yoga nidra intervention, though the Mr2 recording did not show any significant change in scores as compared to baseline. On further analysis it was evident that yoga nidra intervention only showed deterioration in neutral stimuli recognition without affecting the emotion recognition of happy stimuli and in fact improved recognition of anger and fear stimuli. The exact mechanism of this cannot be understood and would require further study. The relative improvement in non-neutral emotional recognition task, though not significant, is also interesting and requires more deliberation with special emphasis on emotion recognition and thus social interaction.

The study highlights the important effect of yoga nidra practice on the cognitive processing. Though there are limitations of this study as it demonstrates effects of yoga nidra practice for two weeks and the EEG locations studied while doing the cognitive tasks were also limited, results with larger samples and longer durations may give a better perspective. This study definitely opens up an opportunity for the use of an easy to do practice of yoga nidra for population health using standardized supervised model. It also highlights the possible role of this practice in promoting learning and memory in healthy subjects and might hold promise in future studies for patients especially with mild learning disability or with mild cognition disorder.

In the post pandemic times, sleep is commonly found to be affected which creates a risk of multiple disorders including neuropsychiatric disorders^22^. This practice for insomnia problem along with awareness of physicians even at the primary care level about simple to do sleep practice guidelines and sleep management thus may be of value ^23^.

## Supporting information

Supplementary Figure 1 (a-j)-Trend line graphs of average accuracy for Cognition Test Battery

Supplementary Figure 1 (a-j)-Trend line graphs of average Reaction Time for Cognition Test Battery

## Data Availability

All data produced in the present study are available upon reasonable request to the authors

## Acknowledgement

Authors acknowledge Department of Science and Technology, Government of India for financial support vide Reference No. 28 May 2020 under Science and Technology for Yoga and Meditation (SATYAM) to carry out this work. Authors also thank the subjects and staff for the support in conducting the study.

## Declaration of Interests

We declare no competing interest.

## Data Sharing

Original data in the coded form (unidentified), including statistical analysis, software used and codes will be made freely available after publication upon request to the corresponding author.

## Notes

### Competing Interest Statement

The authors have declared no competing interest.

### Clinical Trial

Clinical Trials Registry of India- CTRI/2021/02/031349

### Clinical Protocols

https://ctri.nic.in/Clinicaltrials/pmaindet2.php?trialid=52487&EncHid=&userName=Karuna%20Datta

### Funding Statement

Department of Science and Technology SATYAM, Govt of India, sanction letter No. 26 May 2020

### Author Declarations

Ethics Committee of Armed Forces Medical College, Pune, India approved the work (IEC/2019/255 dated 25 Apr 2019). Study registered in Clinical Trials Registry of India- CTRI/2021/02/031349

