## Supplemental Table 1 for "Improved Sleep, Cognitive Processing and Enhanced Learning and Memory Task Accuracy with Yoga Nidra Practice in Novices"

**Supplementary Table 1: ERT- Emotion wise analysis of accuracy score measure in subjects before (baseline) and after (post-intervention) yoga nidra training in test condition comparison and time wise comparison**

| **Sr.**  **No.** | **Emotion** | **Time/Test Condition** | **RM-ANOVA**  **(p-value)** | **Accuracy**  **(Mean ± SD)** | **Post-Hoc**  **(P-value)** | **Difference***  **Mean ± SD** | **95% CI Difference*** | **%Change** |
| --- | --- | --- | --- | --- | --- | --- | --- | --- |
| 1· | Fear | Mr1 | ns |  |  |  |  |  |
|  |  | Mr2 | ns |  |  |  |  |  |
|  |  | PL | ns |  |  |  |  |  |
|  | Fear | CEO | 0·010 | Mr1(0‧449±0·228)  PL(0·582±0·165) | 0·017 | 0·133 ± 0·230 | (0·042, 0·224) | 29·621% |
|  |  | CEC | 0·001 | Mr1(0·426±0·191)  Mr2(0·597±0·212) | 0·001 | 0·171 ± 0·222 | (0·083, 0·259) | 40·141% |
|  |  |  |  | Mr1(0·426±0·191)  PL(0·549±0·182) | 0·042 | 0·123 ± 0·243 | (0·027, 0·219) | 28·873% |
|  |  | YN1W | ns |  |  |  |  |  |
|  |  | YN2W | ns |  |  |  |  |  |
| 2· | Happy | Mr1 | ns |  |  |  |  |  |
|  |  | Mr2 | ns |  |  |  |  |  |
|  |  | PL | ns |  |  |  |  |  |
|  | Happy | CEO | ns |  |  |  |  |  |
|  |  | CEC | ns |  |  |  |  |  |
|  |  | YN1W | ns |  |  |  |  |  |
|  |  | YN2W | ns |  |  |  |  |  |
| 3· | Sad | Mr1 | ns |  |  |  |  |  |
|  |  | Mr2 | ns |  |  |  |  |  |
|  |  | PL | ns |  |  |  |  |  |
|  | Sad | CEO | 0·001 | Mr1(0·653±0·178)  Mr2(0·458±0·233) | 0·0003 | -0·194 ± 0·217 | (-0·280, -0·108) | -29·862% |
|  |  | CEC | 0·000 | Mr1(0·657±0·198)  Mr2(0·50±0·166) | 0·0002 | -0·157 ± 0·171 | (-0·225, -0·089) | -23·896% |
|  |  |  |  | Mr1(0·657±0·198)  PL(0·521±0·219) | 0·003 | -0·136 ± 0·188 | (-0·211, -0·062) | -20·700% |
|  |  | YN1W | ns |  |  |  |  |  |
|  |  | YN2W | ns |  |  |  |  |  |
| 4· | Anger | Mr1 | ns |  |  |  |  |  |
|  |  | Mr2 | ns |  |  |  |  |  |
|  |  | PL | 0·003 | CEO(0·340±0·182)  YN2W(0·495±0·204) | 0·004 | 0·155 ± 0·206 | (0·074, 0·237) | 45·588% |
|  | Anger | CEO | 0·000 | Mr1(0·481±0·172)  PL(0·340±0·182) | 0·004 | -0·141 ± 0·205 | (-0·222, -0·060) | -29·314% |
|  |  |  |  | Mr2(0·537±0·202)  PL(0·340±0·182) | 0·002 | -0·197 ± 0·260 | (-0·301, -0·092) | -36·685% |
|  |  | CEC | 0·002 | Mr1(0·505±0·175)  PL(0·388±0·191) | 0·008 | -0·116 ± 0·183 | (-0·189, -0·044) | -5·909% |
|  |  |  |  | Mr2(0·495±0·145)  PL(0·388±0·191) | 0·009 | -0·107 ± 0·169 | (-0·174, -0·040) | -5·297% |
|  |  | YN1W | 0·018 | Mr2(0·505±0·185)  PL(0·380±0·185) | 0·048 | -0·125 ± 0·252 | (-0·224, -0·025) | -24·752% |
|  |  | YN2W | ns |  |  |  |  |  |
| 5· | Neutral | Mr1 | 0·004 | CEC(0·593±0·285)  YN2W(0·394±0·289) | 0·010 | -0·199 ± 0·267 | (-0·305, -0·093) | -11·801% |
|  |  |  |  | CEO(0·569±0·226)  YN2W(0·394±0·289) | 0·036 | -0·175± 0·313 | (-0·299, -0·052) | -30·756% |
|  |  | Mr2 | ns |  |  |  |  |  |
|  |  | PL | ns |  |  |  |  |  |
|  | Neutral | CEO | 0·003 | Mr1(0·569±0·226)  Mr2(0·449±0·260) | 0·009 | -0·120 ± 0·178 | (-0·191, -0·050) | -6·828% |
|  |  | CEC | 0·037 | Mr1(0·592±0·285)  Mr2(0·468±0·251) | 0·012 | -0·125 ± 0·196 | (-0·203, -0·047) | -20·946% |
|  |  | YN1W | ns |  |  |  |  |  |
|  |  | YN2W | ns |  |  |  |  |  |

Notes: Test conditions- CEO: Control with Eyes Open, CEC: Control with Eyes Close, YN1W: At the end of one week of Yoga-nidra practice after training, YN2W: At the end of two weeks of Yoga-nidra practice after training·

Time- Mr1: Test just before CEC, CEO, YN1W or YN2W, Mr2: Tests conducted just after CEO, CEC, YN1W or YN2W, PL: Tests conducted after the lunch·

RM-ANOVA: One way repeated measure ANOVA was used for checking significant change (given p-values)·

Post-Hoc: Post-hoc analysis (pairwise comparison) is done using Wilcoxon signed rank test with Bonferroni correction·

ns: not significant

*Difference is calculated as difference between two accuracy scores given in Accuracy column (i.e. 5^th^)
