## Supplementary Table 2 for "Improved Sleep, Cognitive Processing and Enhanced Learning and Memory Task Accuracy with Yoga Nidra Practice in Novices"

**Supplementary Table 2: Cognitive battery testing (CBT) parameters (Mean ± SD) of all cognitive battery tests in the subjects**

| Sr.  No | **Test Time** | **Time**  **Test**  **Condition** | **Mr1** | | | | **Mr2** | | | | **PL** | | | |
| --- | --- | --- | --- | --- | --- | --- | --- | --- | --- | --- | --- | --- | --- | --- |
|  | **CBT**  **(Cognitive**  **Domains Assessed)** | **CBT**  **Parameter** | **CEC** | **CEO** | **YNW1** | **YNW2** | **CEC** | **CEO** | **YNW1** | **YNW2** | **CEC** | **CEO** | **YN1W** | **YN2W** |
| 1 | **MPT**  **(Sensory-motor speed)** | Reaction time | 0·531±0·293 | 0·724±0·855 | 0·460±0·164 | 0·456±0·178 | 0·507±0·218 | 0·50±0·295 | 0·473±0·196 | 0·462±0·188 | 0·492±0·197 | 0·495±0·201 | 0·462±0·192 | 0·482±0·263 |
|  |  | Accuracy | 0·703±0·162 | 0·675±0·205 | 0·694±0·173 | 0·687±0·170 | 0·709±0·168 | 0·713±0·166 | 0·707±0·168 | 0·689±0.177 | 0·711±0·166 | 0·729±0·156 | 0·717±0·165 | 0·695±0·163 |
| 2 | **VOLT**  **(Spatial learning and memory)** | Reaction time | 2·193±1·607 | 2·676±2·970 | 1·624±1·184 | 1·479±0·972 | 1·758±1·277 | 2·078±1·796 | 1·591±1·164 | 1·443±1·049 | 1·705±1·181 | 1·839±1·466 | 1·538±1·202 | 1·346 ±1·034 |
|  |  | Accuracy | 0·733±0·097 | 0·665±0·148 | 0·781±0·102 | 0·775±0·112 | 0·757±0·136 | 0·718±0·131 | 0·781±0·116 | 0·764±0·134 | 0·745±0·140 | 0·702±0·159 | 0·770±0·143 | 0·748±0·149 |
| 3 | **NBACK**  **(Working memory)** | Reaction time | 1·334±0·612 | 1·296±0·626 | 1·352±0·581 | 1·323±0·598 | 1·312±0·616 | 1·294±0·619 | 1·366±0·576 | 1·341±0·590 | 1·288±0·633 | 1·30±0·609 | 1·36±0·580 | 1·344±0·594 |
|  |  | Accuracy | 0·674±0·165 | 0·668±0·172 | 0·732±0·148 | 0·749±0·153 | 0·716±0·173 | 0·708±0·152 | 0·783±0·141 | 0·760±0·139 | 0·707±0·193 | 0·713±0·176 | 0·747±0·142 | 0·750±0·145 |
| 4 | **AIM**  **(Abstraction, concept formation)** | Reaction time | 2·381±1·821 | 2·923±2·409 | 2·166±1·745 | 1·795±1·624 | 2·334±1·921 | 2·262±1·796 | 2·271±2·188 | 1·559±1·360 | 2·268±2·036 | 1·983±1·541 | 1·734±1·603 | 1·594 ±1·542 |
|  |  | Accuracy | 0·554±0·093 | 0·509±0·09 | 0·560±0·118 | 0·584±0·111 | 0·515±0·126 | 0·546±0·123 | 0·567±0·115 | 0·569±0·140 | 0·521±0·146 | 0·547±0·134 | 0·565±0·143 | 0·578±0·133 |
| 5 | **LOT**  **(Spatial orientation)** | Reaction time | 8·501±4·652 | 9·098±5·296 | 7·707±4·763 | 6·712±3·937 | 7·869±5·243 | 7·628±4·672 | 7·509±5·055 | 6·862±4·459 | 7·281±4·884 | 7·554±4·447 | 6·861±4·258 | 6·469±4·539 |
|  |  | Accuracy | 0·664±0·206 | 0·652±0·157 | 0·665±0·132 | 0·704±0·128 | 0·654±0·225 | 0·678±0·124 | 0·672±0·160 | 0·658±0·172 | 0·665±0·173 | 0·698±0·141 | 0·725±0·088 | 0·656±0·174 |
| 6 | **ERT**  **(Emotion identification)** | Reaction time | 2·556±2·490 | 3·227±3·989 | 2·325±2·245 | 2·14±2·124 | 2·16±1·987 | 2·243±2·443 | 2·076±1·877 | 1·82±1·791 | 2·207±2·505 | 2·225±2·538 | 1·908±1·535 | 1·965±2·167 |
|  |  | Accuracy | 0·620±0·071 | 0·619±0·080 | 0·590±0·078 | 0·559±0·076 | 0·596±0·097 | 0·590±0·102 | 0·591±0·103 | 0·558±0·102 | 0·577±0·093 | 0·570±0·088 | 0·581±0·081 | 0·569±0·078 |
| 7 | **MRT**  **(Abstract reasoning)** | Reaction time | 11·90±7·996 | 12·405±8·290 | 11·80±8·24 | 10·236±7·24 | 11·015±7·955 | 11·06±8·044 | 11·014±7·884 | 10·665±7·965 | 10·305±7·689 | 10·891±7·871 | 10·192±7·297 | 10·665±7·909 |
|  |  | Accuracy | 0·630±0·157 | 0·630±0·202 | 0·657±0·162 | 0·688±0·154 | 0·611±0·142 | 0·580±0·187 | 0·645±0·196 | 0·651±0·167 | 0·665±0·158 | 0·642±0·209 | 0·704±0·175 | 0·676±0·159 |
| 8 | **DSST**  **(Complex scanning and visual tracking)** | Reaction time | 1·135±0·548 | 1·199±0·766 | 1·097±0·532 | 1·051±0·453 | 1·105±0·401 | 1·114±0·482 | 1·092±0·470 | 1·094±0·549 | 1·077±0·406 | 1·077±0·426 | 1·068±0·464 | 1·049±0·439 |
|  |  | Accuracy | 0·990±0·019 | 0·987±0·023 | 0·991±0·013 | 0·977±0·093 | 0·990±0·015 | 0·990±0·019 | 0·994±0·014 | 0·989±0·018 | 0·991±0·015 | 0·985±0·040 | 0·991±0·012 | 0·990±0·017 |
| 9 | **BART**  **(Risk decision making)** | Reaction time | 0·674±0·764 | 0·743±0·780 | 0·590±0·646 | 0·477±0·573 | 0·611±0·657 | 0·603±0·638 | 0·492±0·537 | 0·412±0·429 | 0·586±0·699 | 0·581±0·581 | 0·435±0·527 | 0·387±0·399 |
|  |  | Adjusted number of pumps | 4·144±1·035 | 4·160±1·198 | 4·284±0·959 | 4·223±0·955 | 4·167±1·021 | 4·183±1·110 | 4·171±1·058 | 4·431±0·954 | 3·730±1·300 | 3·940±0·797 | 4·172±1·130 | 4·576±0·994 |
|  |  | Cash collected($) | 92·519±15·575 | 90·630±16·727 | 91±14·167 | 96·37±13·545 | 83·333±11·655 | 79·259±12·687 | 78·889±10·984 | 83·185±19·472 | 78·667±17·746 | 77±15·161 | 83·296±13·575 | 83·852±16·221 |
|  |  | Number of balloons burst | 11·111±2·407 | 11·296±3·232 | 11·815±3·039 | 11·037±3·216 | 13·222±3·13 | 13·556±3·238 | 13·63±3·477 | 14·185±3·981 | 12·296±3·441 | 13·889±3·378 | 12·889±3·534 | 14·593±3·422 |
|  |  | Risk Propensity | 6·062±1·885 | 6·39±2·601 | 6·39±2·17 | 6·254±2·042 | 6·590±2·16 | 6·838±2·981 | 6·7±2·701 | 8·356±6·679 | 6·120±2·346 | 6·828±2·915 | 6·742±3·111 | 8·081±2·783 |
| 10 | **PVT**  **(Vigilant attention)** | Reaction time | 0·302±0·408 | 0·397±0·878 | 0·339±0·502 | 0·306±0·334 | 0·29±0·286 | 0·345±0·754 | 0·318±0·351 | 0·297±0·317 | 0·283±0·247 | 0·326±0·642 | 0·276±0·267 | 0·302 ±0·392 |
|  |  | Accuracy | 0·784±0·178 | 0·728±0·255 | 0·803±0·161 | 0·838±0·111 | 0·848±0·084 | 0·793±0·193 | 0·840±0·150 | 0·827±0·121 | 0·815±0·097 | 0·825±0·139 | 0·822±0·140 | 0·830±0·140 |
|  |  | Number of false starts | 13·2±11·0 | 16·0±16·9 | 9·67±8·55 | 8·67±9·76 | 8·22±7·29 | 12·4±12·9 | 7·26±6·84 | 9·70±11·8 | 11·1±11·1 | 10·1±9·06 | 12·1±12·7 | 10·6±17·0 |

Cognitive battery Tests (CBT) - MPT: Motor Praxis Task, VOLT: Visual Object Learning Task, NBACK: Fractal 2-Back, AIM: Abstract Matching, LOT: Line Orientation Task, ERT: Emotion Recognition Task, MRT: Matrix Reasoning Task, DSST: Digital Symbol Substitution Task, PVT: Psychomotor Vigilance Task, BART: Balloon Analog Risk Task

Test Conditions - CEO: Control with Eyes Open, CEC: Control with Eyes Close, YN1W: At the end of one week of Yoga-nidra practice after training, YN2W: At the end of two weeks of Yoga-nidra practice after training·

Times - Mr1: Test just before CEC, CEO, YN1W or YN2W, Mr2: Tests conducted just after CEO, CEC, YN1W or YN2W, PL: Tests conducted after the lunch
