## Supplementary Figure 1 (a-j)-Trend line graphs of average accuracy for Cognition Test Battery for "Improved Sleep, Cognitive Processing and Enhanced Learning and Memory Task Accuracy with Yoga Nidra Practice in Novices"

### Slide 1
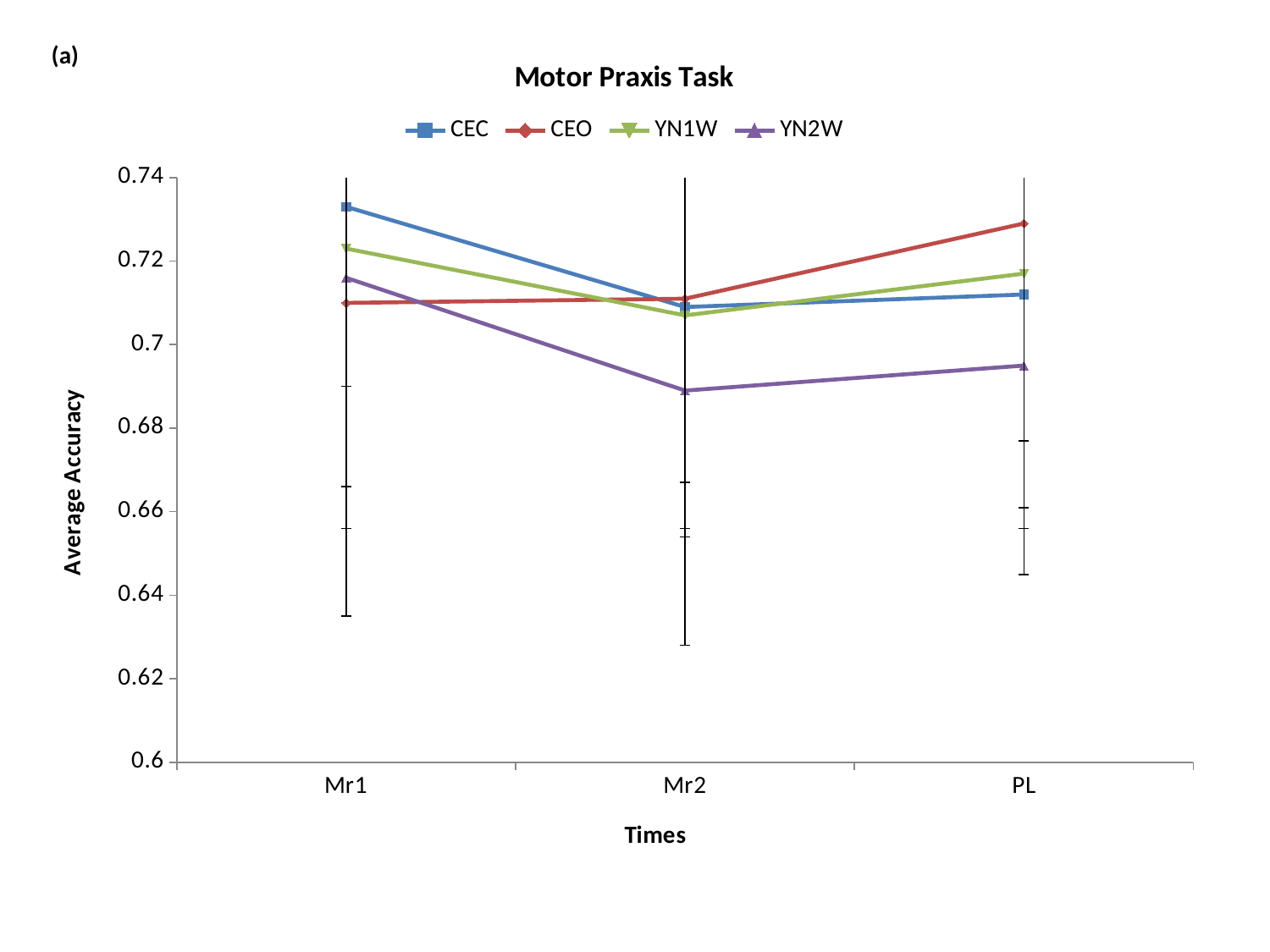

#### Chart: Motor Praxis Task
| Category | | | | |
|---|---|---|---|---|
| Mr1 | 0.733 | 0.71 | 0.723 | 0.716 |
| Mr2 | 0.709 | 0.711 | 0.707 | 0.689 |
| PL | 0.712 | 0.729 | 0.717 | 0.695 |

### Slide 2
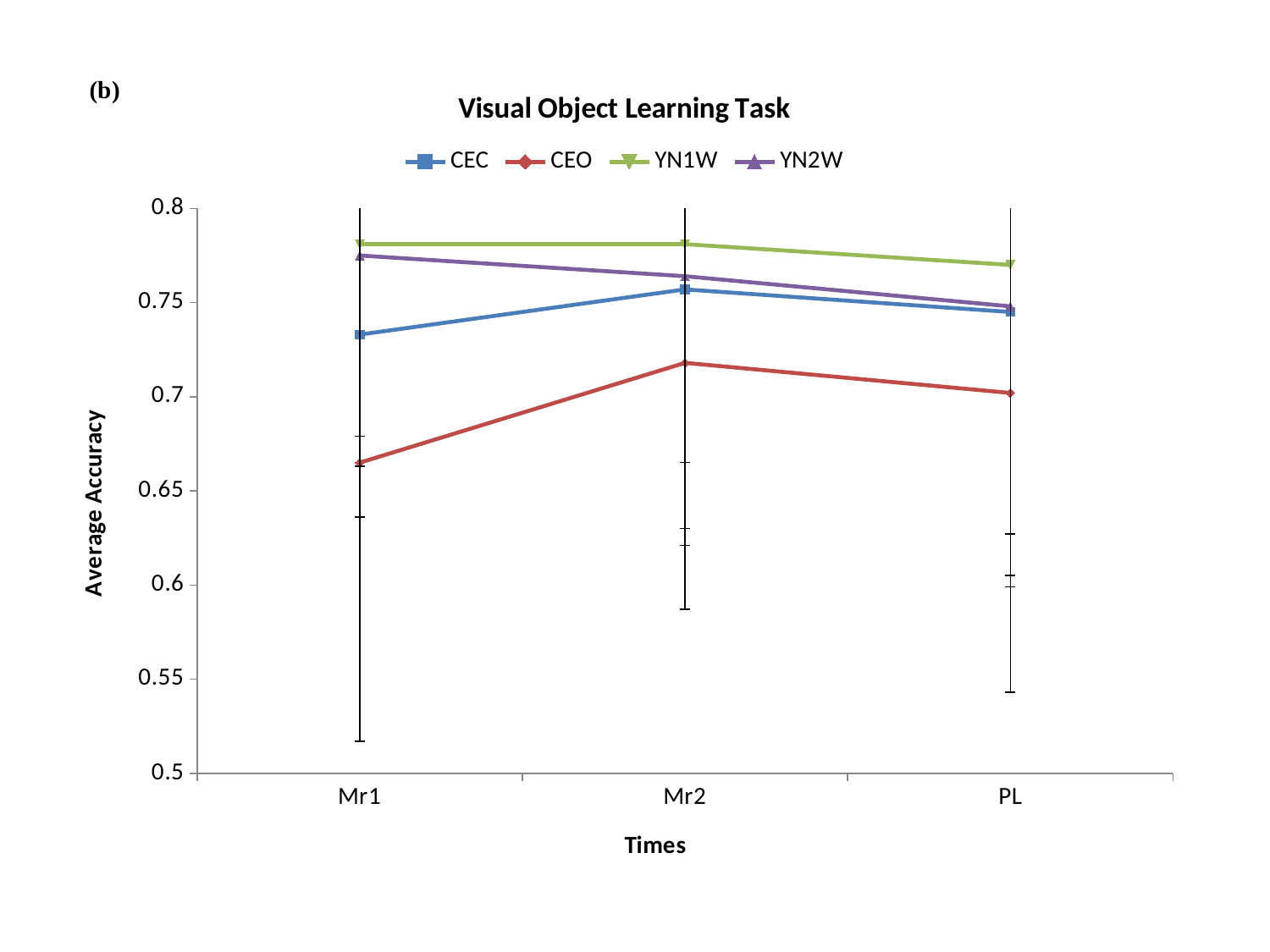

#### Chart: Visual Object Learning Task
| Category | | | | |
|---|---|---|---|---|
| Mr1 | 0.733 | 0.665 | 0.781 | 0.775 |
| Mr2 | 0.757 | 0.718 | 0.781 | 0.764 |
| PL | 0.745 | 0.702 | 0.77 | 0.748 |

### Slide 3
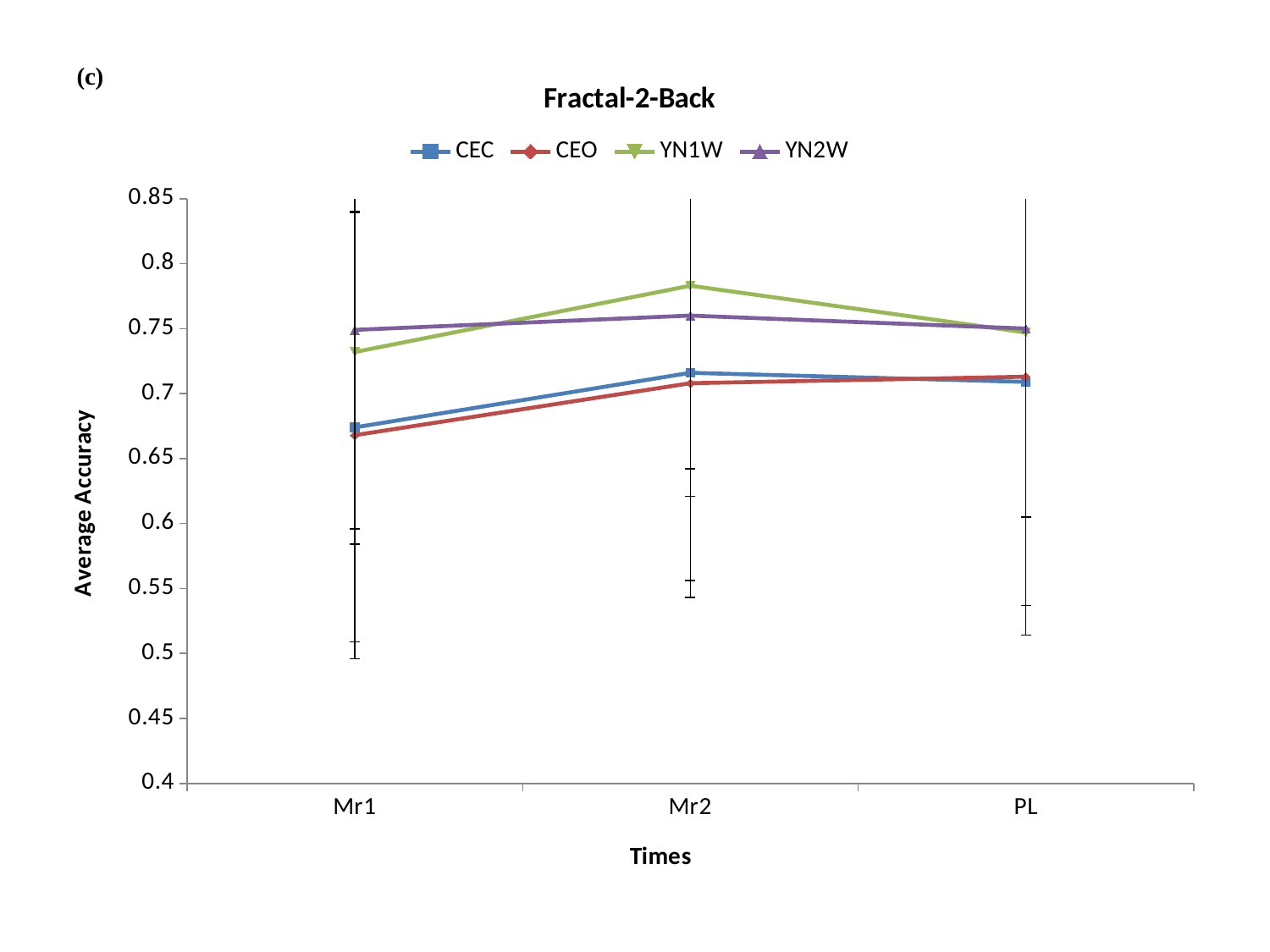

#### Chart: Fractal-2-Back
| Category | | | | |
|---|---|---|---|---|
| Mr1 | 0.674 | 0.668 | 0.732 | 0.749 |
| Mr2 | 0.716 | 0.708 | 0.783 | 0.76 |
| PL | 0.709 | 0.713 | 0.747 | 0.75 |

### Slide 4
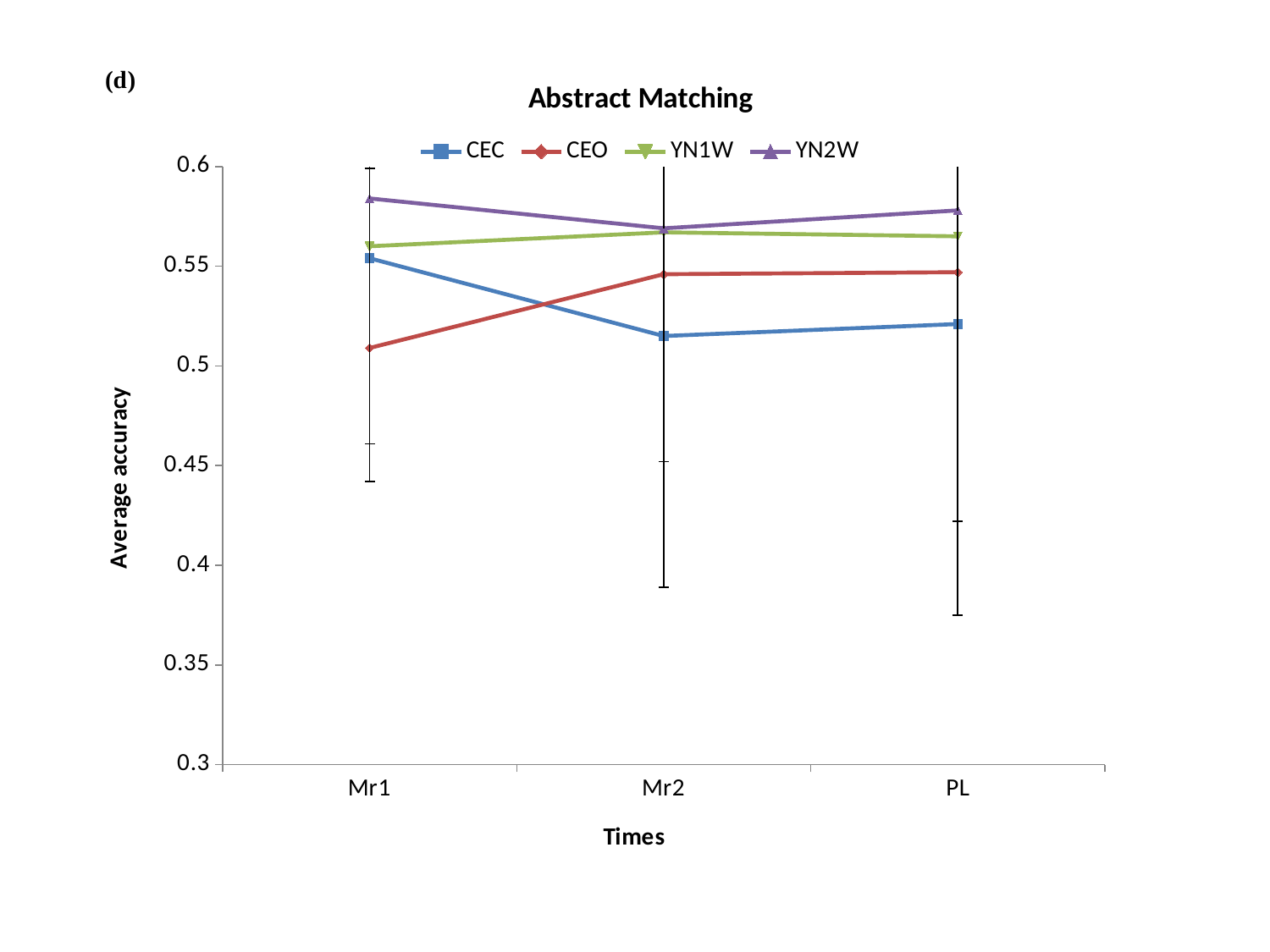

#### Chart: Abstract Matching
| Category | | | | |
|---|---|---|---|---|
| Mr1 | 0.554 | 0.509 | 0.56 | 0.584 |
| Mr2 | 0.515 | 0.546 | 0.567 | 0.569 |
| PL | 0.521 | 0.547 | 0.565 | 0.578 |

### Slide 5
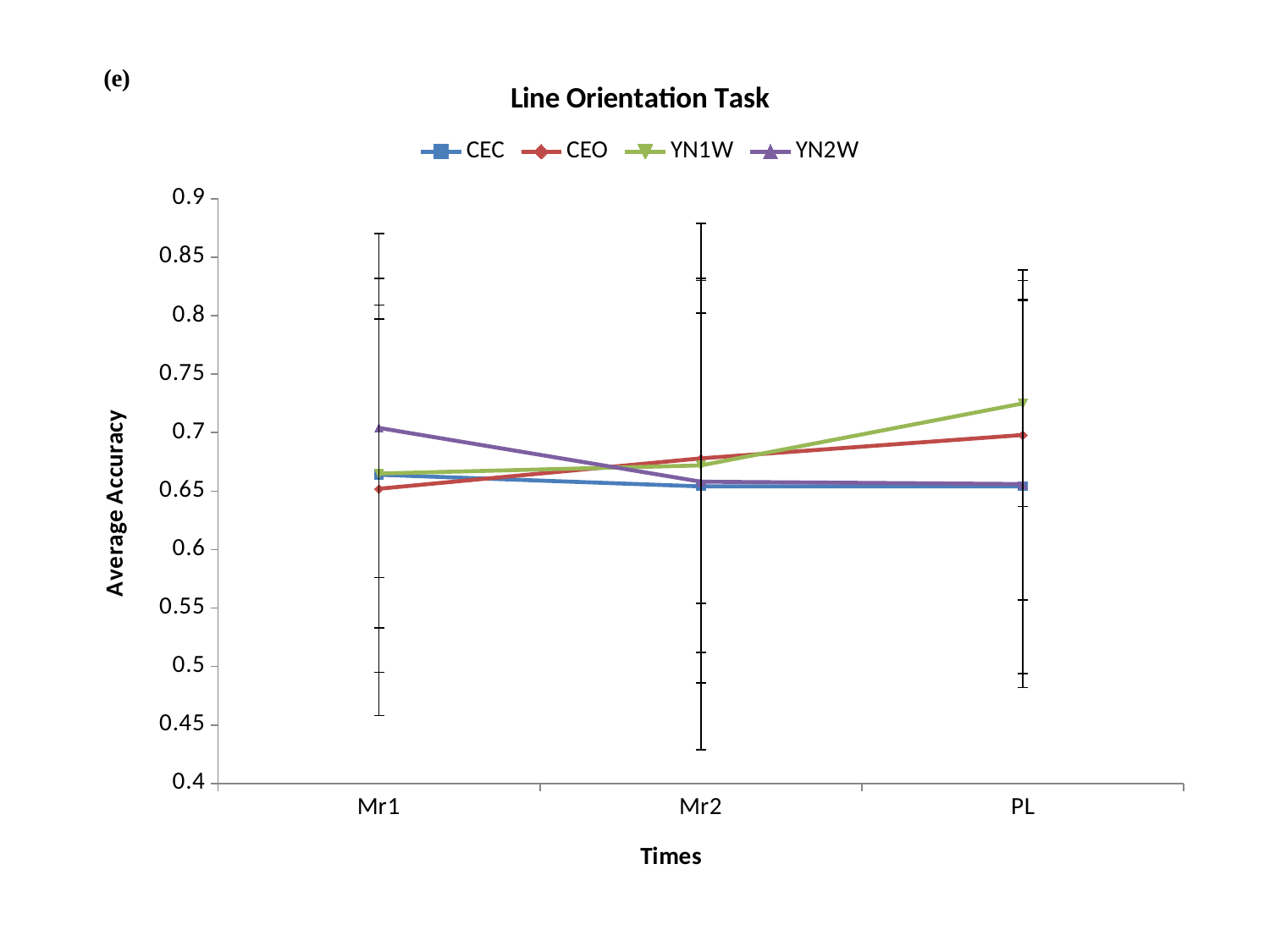

#### Chart: Line Orientation Task
| Category | | | | |
|---|---|---|---|---|
| Mr1 | 0.664 | 0.652 | 0.665 | 0.704 |
| Mr2 | 0.654 | 0.678 | 0.672 | 0.658 |
| PL | 0.654 | 0.698 | 0.725 | 0.656 |

### Slide 6
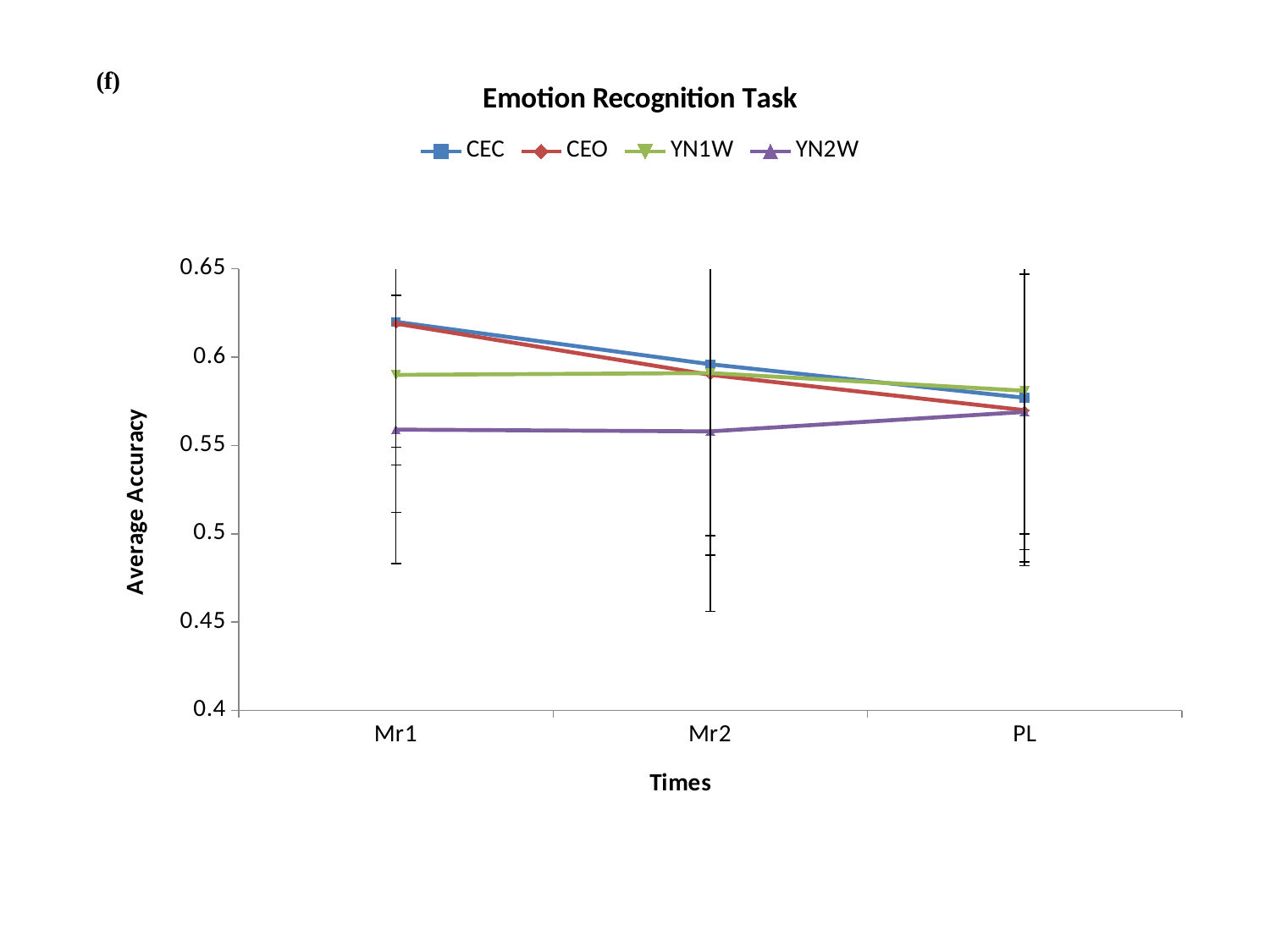

#### Chart: Emotion Recognition Task
| Category | | | | |
|---|---|---|---|---|
| Mr1 | 0.62 | 0.619 | 0.59 | 0.559 |
| Mr2 | 0.596 | 0.59 | 0.591 | 0.558 |
| PL | 0.577 | 0.57 | 0.581 | 0.569 |

### Slide 7
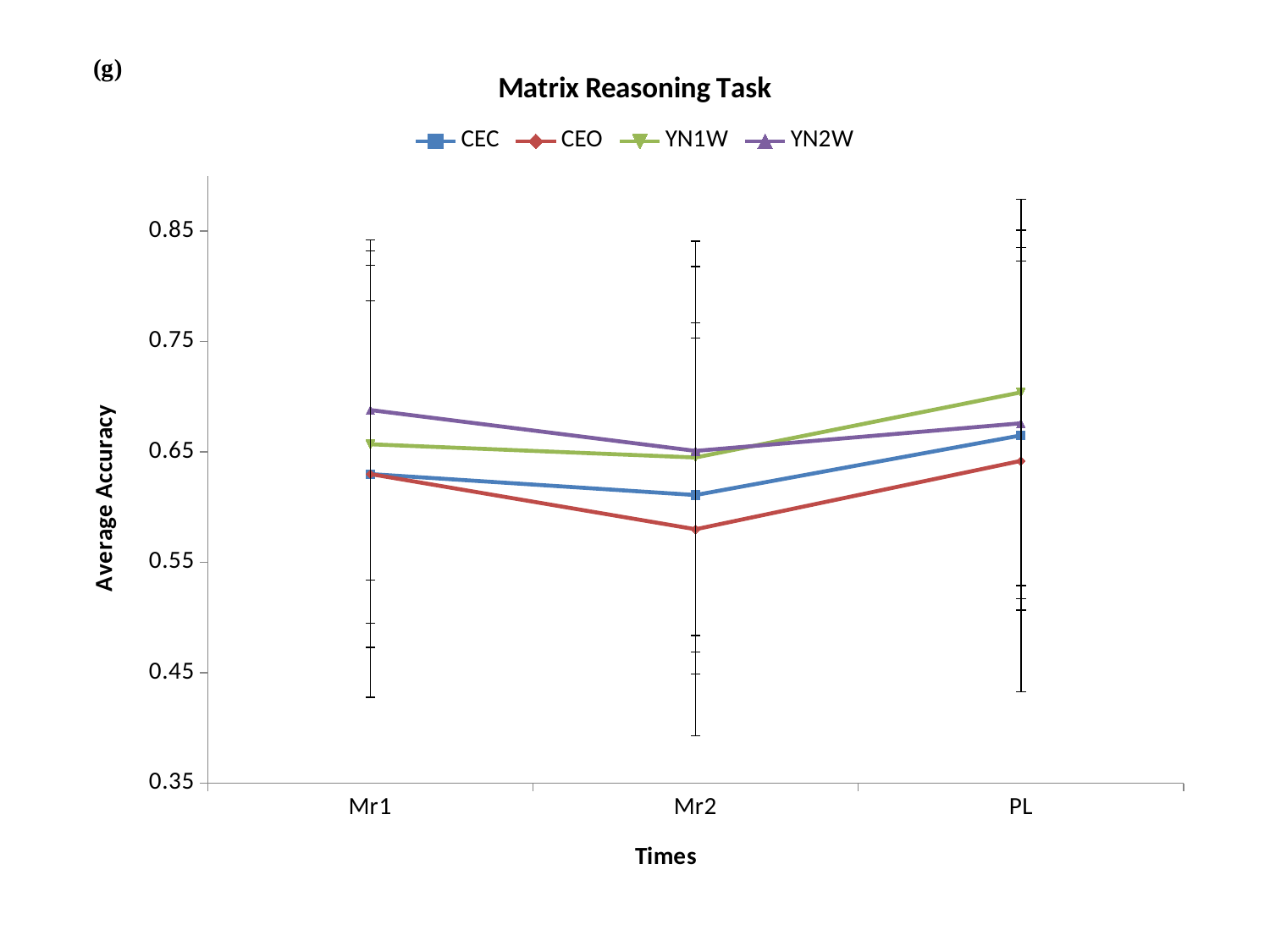

#### Chart: Matrix Reasoning Task
| Category | | | | |
|---|---|---|---|---|
| Mr1 | 0.63 | 0.63 | 0.657 | 0.688 |
| Mr2 | 0.611 | 0.58 | 0.645 | 0.651 |
| PL | 0.665 | 0.642 | 0.704 | 0.676 |

### Slide 8
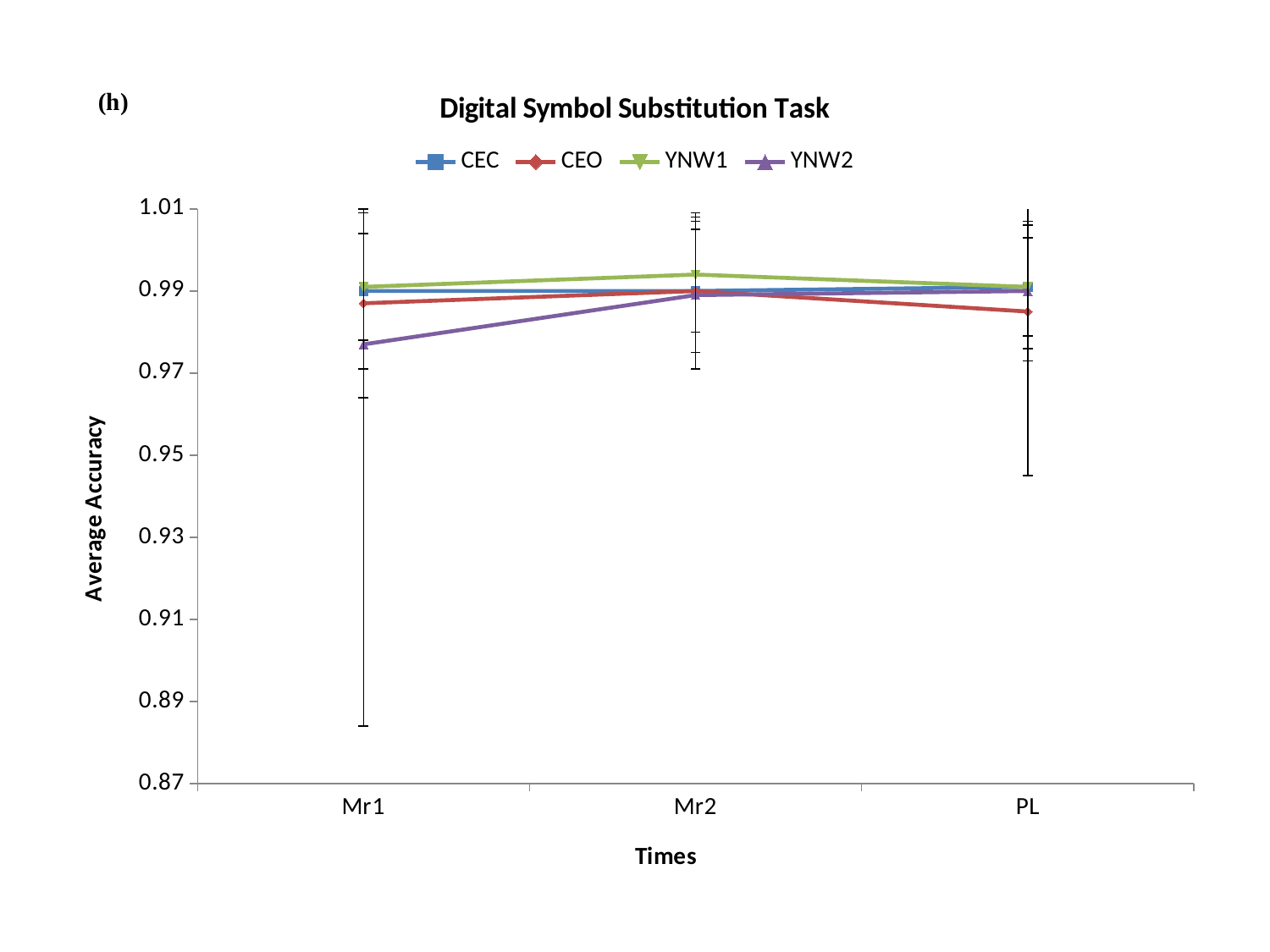

#### Chart: Digital Symbol Substitution Task
| Category | | | | |
|---|---|---|---|---|
| Mr1 | 0.99 | 0.987 | 0.991 | 0.977 |
| Mr2 | 0.99 | 0.99 | 0.994 | 0.989 |
| PL | 0.991 | 0.985 | 0.991 | 0.99 |

### Slide 9
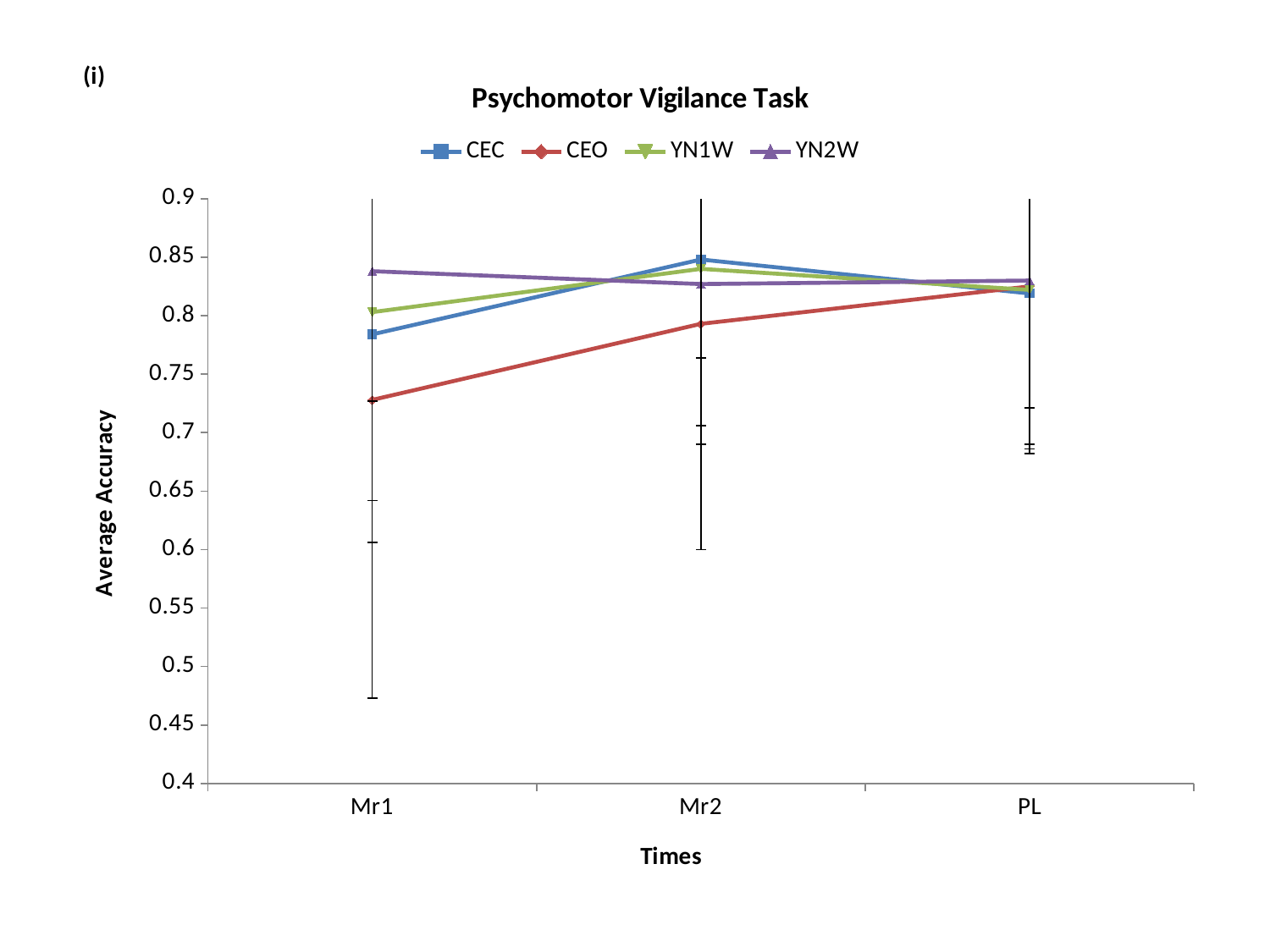

#### Chart: Psychomotor Vigilance Task
| Category | | | | |
|---|---|---|---|---|
| Mr1 | 0.784 | 0.728 | 0.803 | 0.838 |
| Mr2 | 0.848 | 0.793 | 0.84 | 0.827 |
| PL | 0.819 | 0.825 | 0.822 | 0.83 |
