## Supplementary Figure 1 (a-j)-Trend line graphs of average Reaction Time for Cognition Test Battery for "Improved Sleep, Cognitive Processing and Enhanced Learning and Memory Task Accuracy with Yoga Nidra Practice in Novices"

### Slide 1
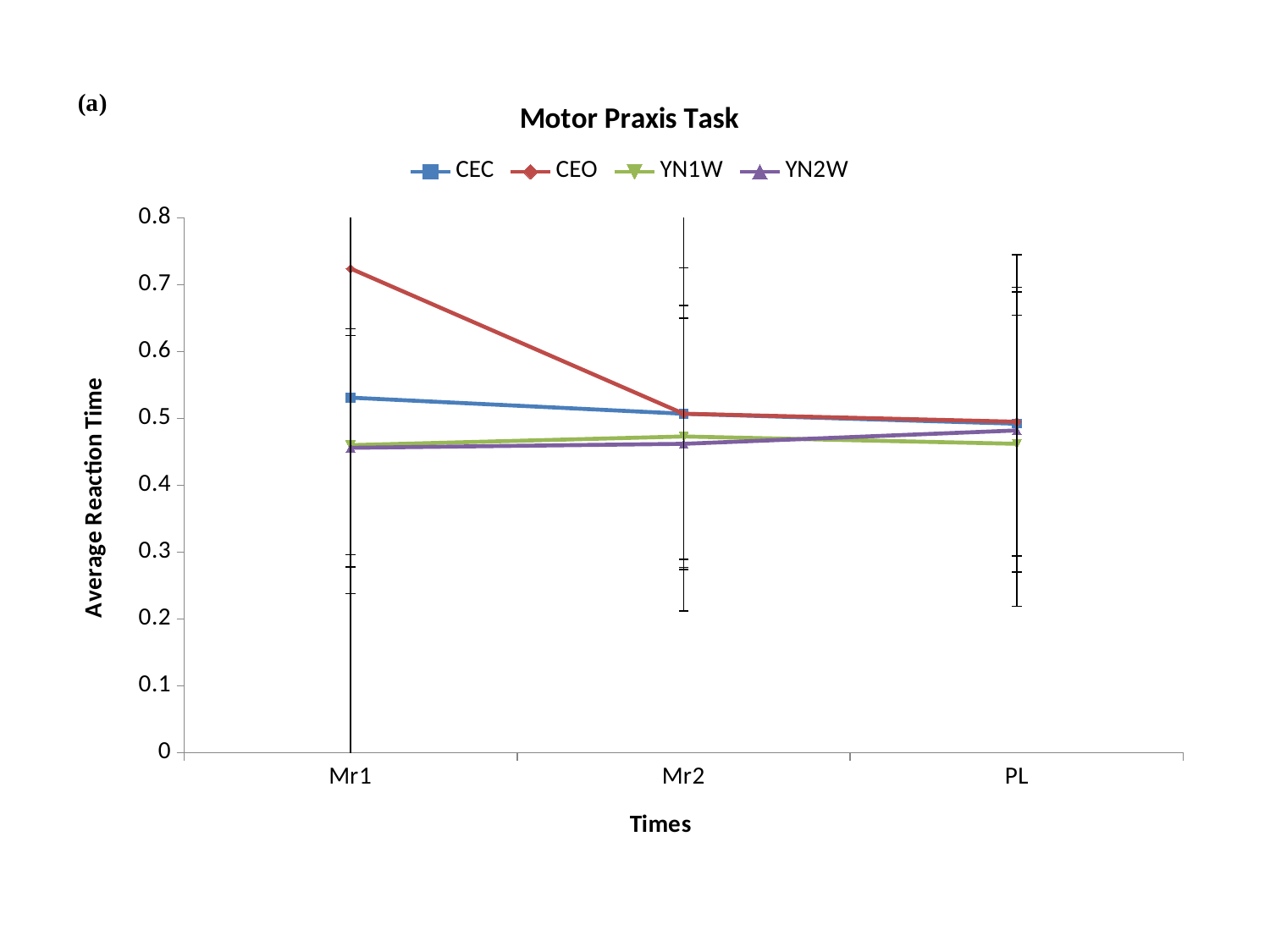

#### Chart: Motor Praxis Task
| Category | CEC | CEO | YN1W | YN2W |
|---|---|---|---|---|
| Mr1 | 0.531 | 0.724 | 0.46 | 0.456 |
| Mr2 | 0.507 | 0.507 | 0.473 | 0.462 |
| PL | 0.492 | 0.495 | 0.462 | 0.482 |

### Slide 2
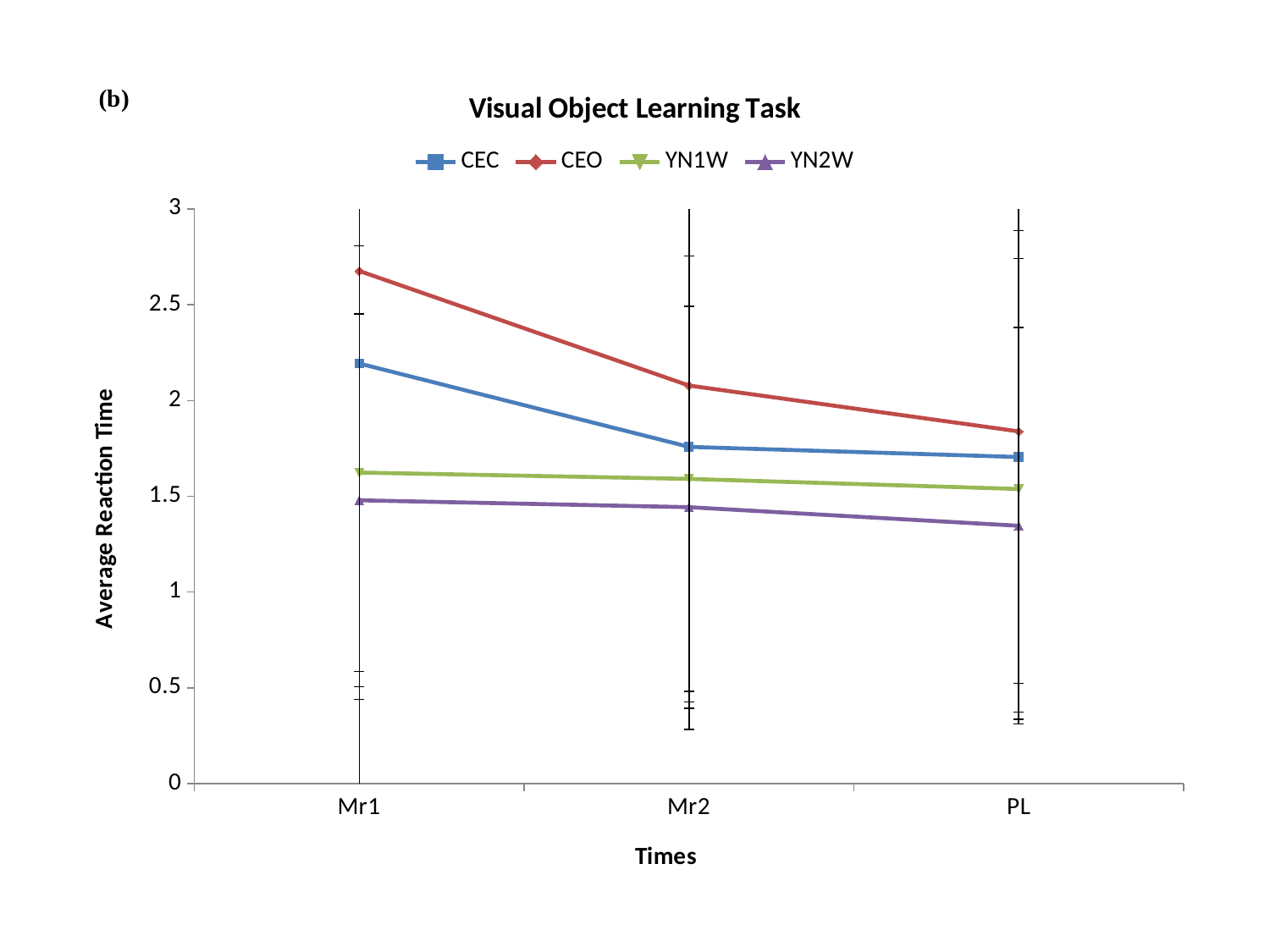

#### Chart: Visual Object Learning Task
| Category | CEC | CEO | YN1W | YN2W |
|---|---|---|---|---|
| Mr1 | 2.193 | 2.676 | 1.624 | 1.479 |
| Mr2 | 1.758 | 2.078 | 1.591 | 1.443 |
| PL | 1.705 | 1.839 | 1.538 | 1.346 |

### Slide 3
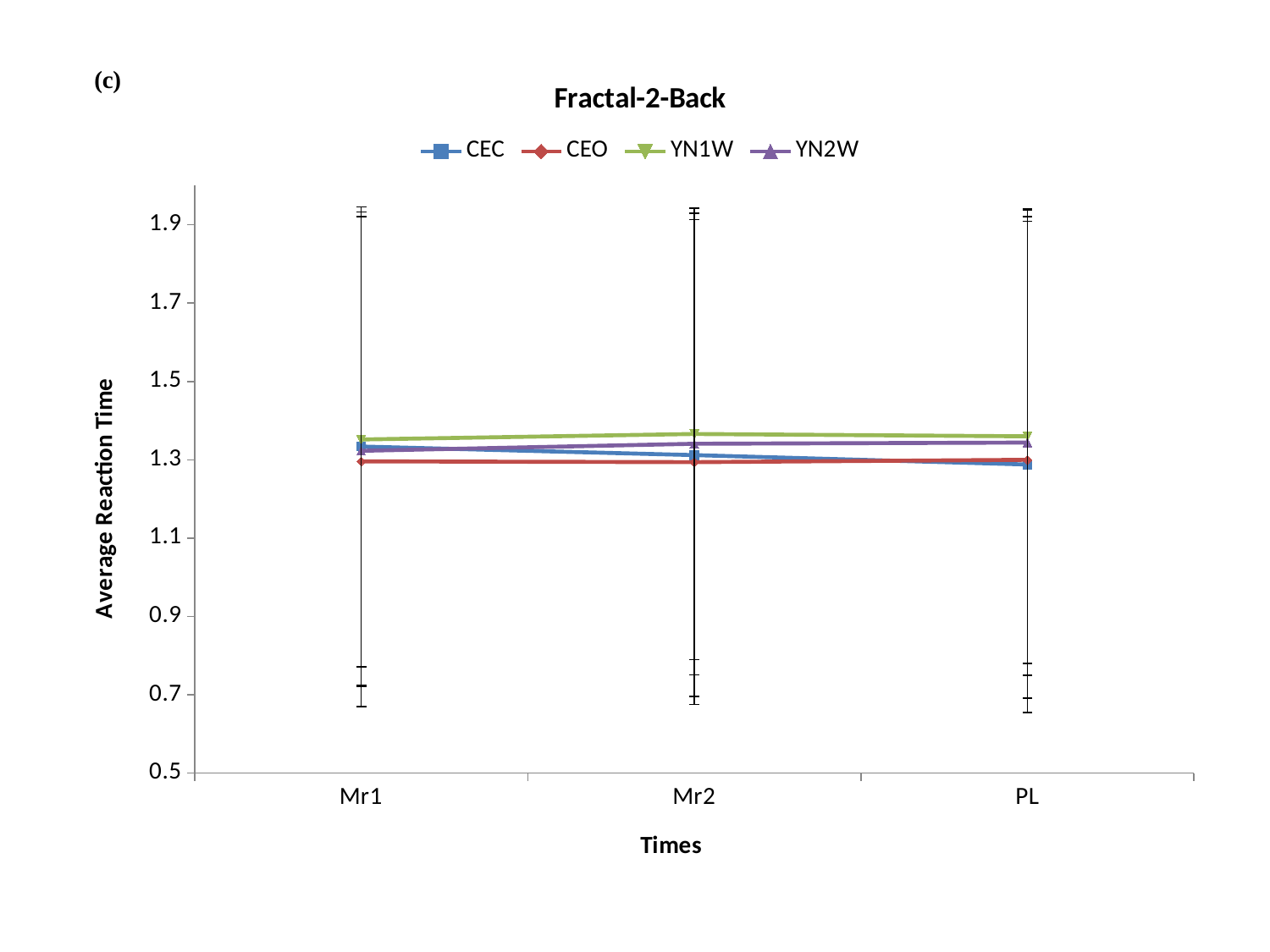

#### Chart: Fractal-2-Back
| Category | CEC | CEO | YN1W | YN2W |
|---|---|---|---|---|
| Mr1 | 1.334 | 1.296 | 1.352 | 1.323 |
| Mr2 | 1.312 | 1.294 | 1.366 | 1.341 |
| PL | 1.288 | 1.3 | 1.36 | 1.344 |

### Slide 4
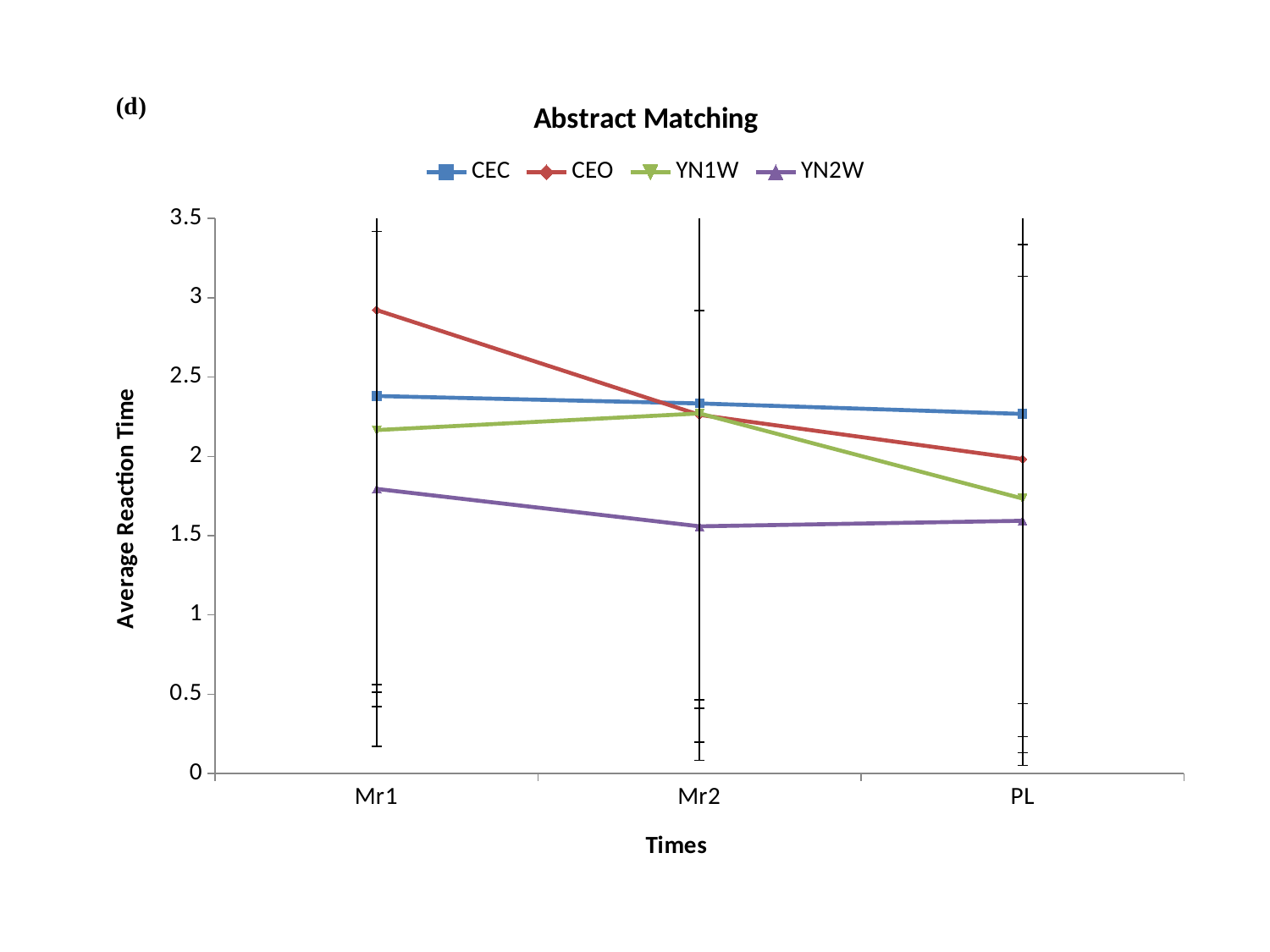

#### Chart: Abstract Matching
| Category | CEC | CEO | YN1W | YN2W |
|---|---|---|---|---|
| Mr1 | 2.381 | 2.923 | 2.166 | 1.795 |
| Mr2 | 2.334 | 2.262 | 2.271 | 1.559 |
| PL | 2.268 | 1.983 | 1.734 | 1.594 |

### Slide 5
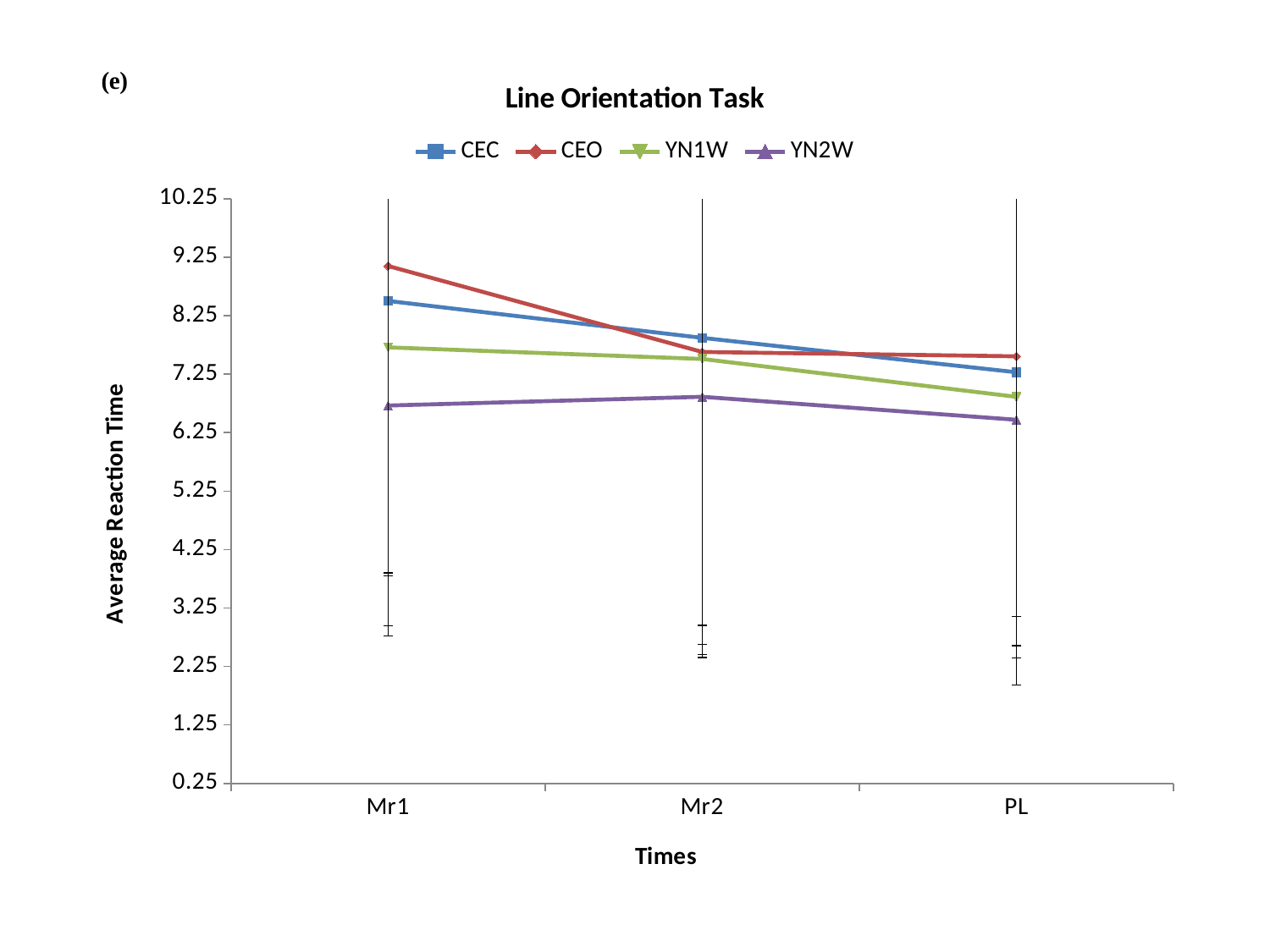

#### Chart: Line Orientation Task
| Category | CEC | CEO | YN1W | YN2W |
|---|---|---|---|---|
| Mr1 | 8.501 | 9.098 | 7.707 | 6.712 |
| Mr2 | 7.869 | 7.628 | 7.509 | 6.862 |
| PL | 7.281 | 7.554 | 6.861 | 6.469 |

### Slide 6
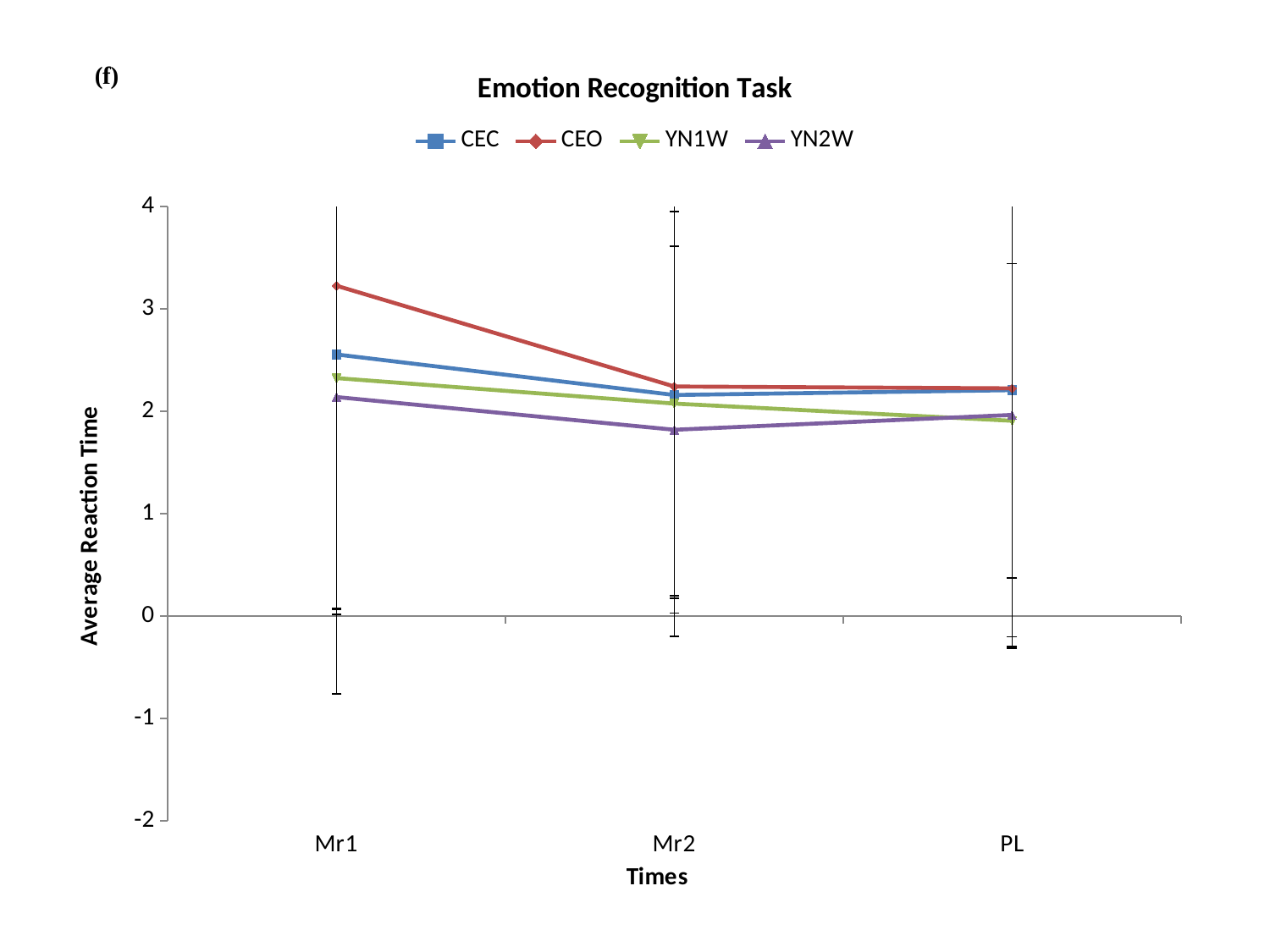

#### Chart: Emotion Recognition Task
| Category | CEC | CEO | YN1W | YN2W |
|---|---|---|---|---|
| Mr1 | 2.556 | 3.227 | 2.325 | 2.14 |
| Mr2 | 2.16 | 2.243 | 2.076 | 1.82 |
| PL | 2.207 | 2.225 | 1.908 | 1.965 |

### Slide 7
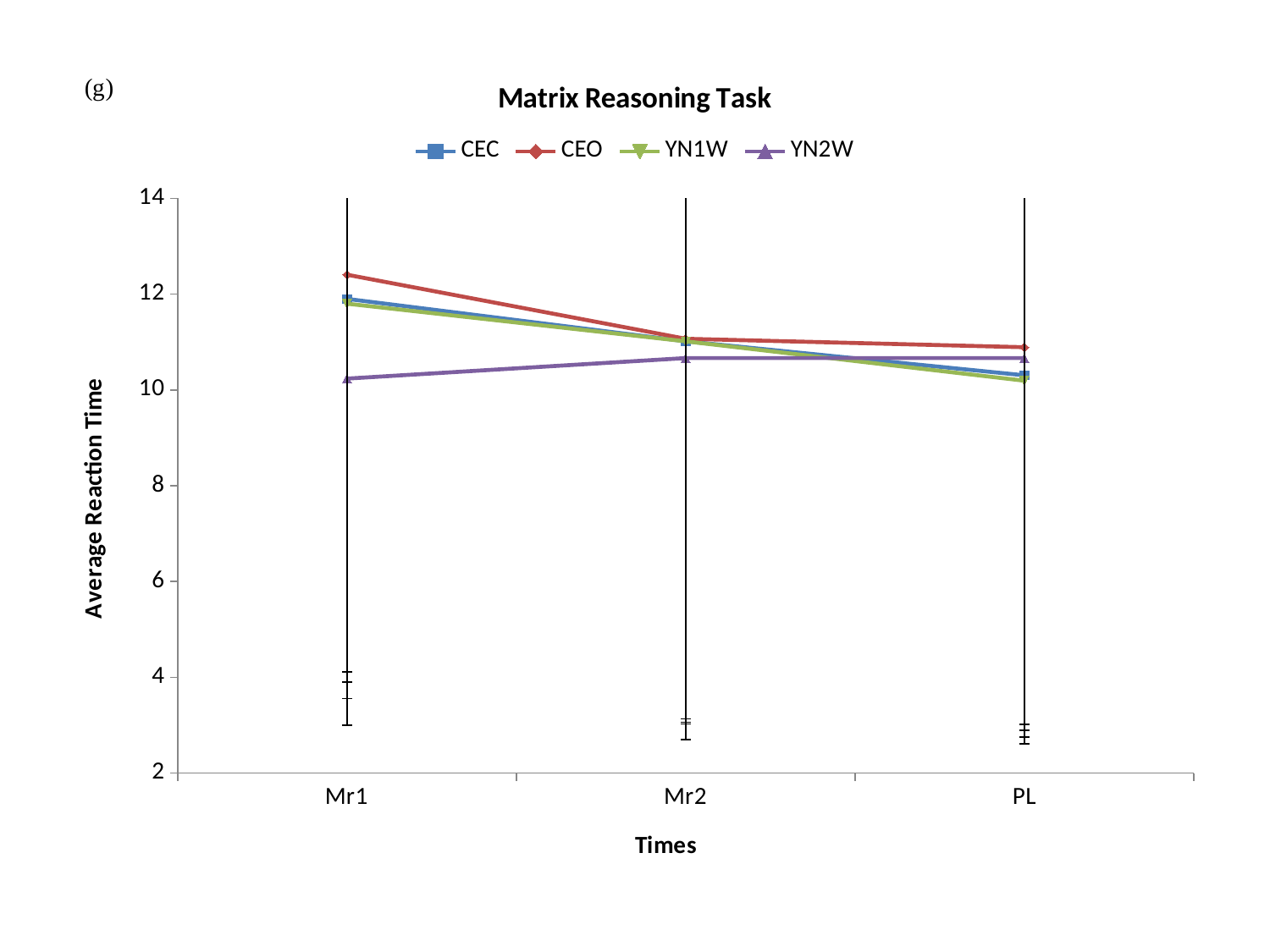

#### Chart: Matrix Reasoning Task
| Category | CEC | CEO | YN1W | YN2W |
|---|---|---|---|---|
| Mr1 | 11.9 | 12.405 | 11.8 | 10.236 |
| Mr2 | 11.015 | 11.067 | 11.014 | 10.665 |
| PL | 10.305 | 10.891 | 10.192 | 10.665 |

### Slide 8
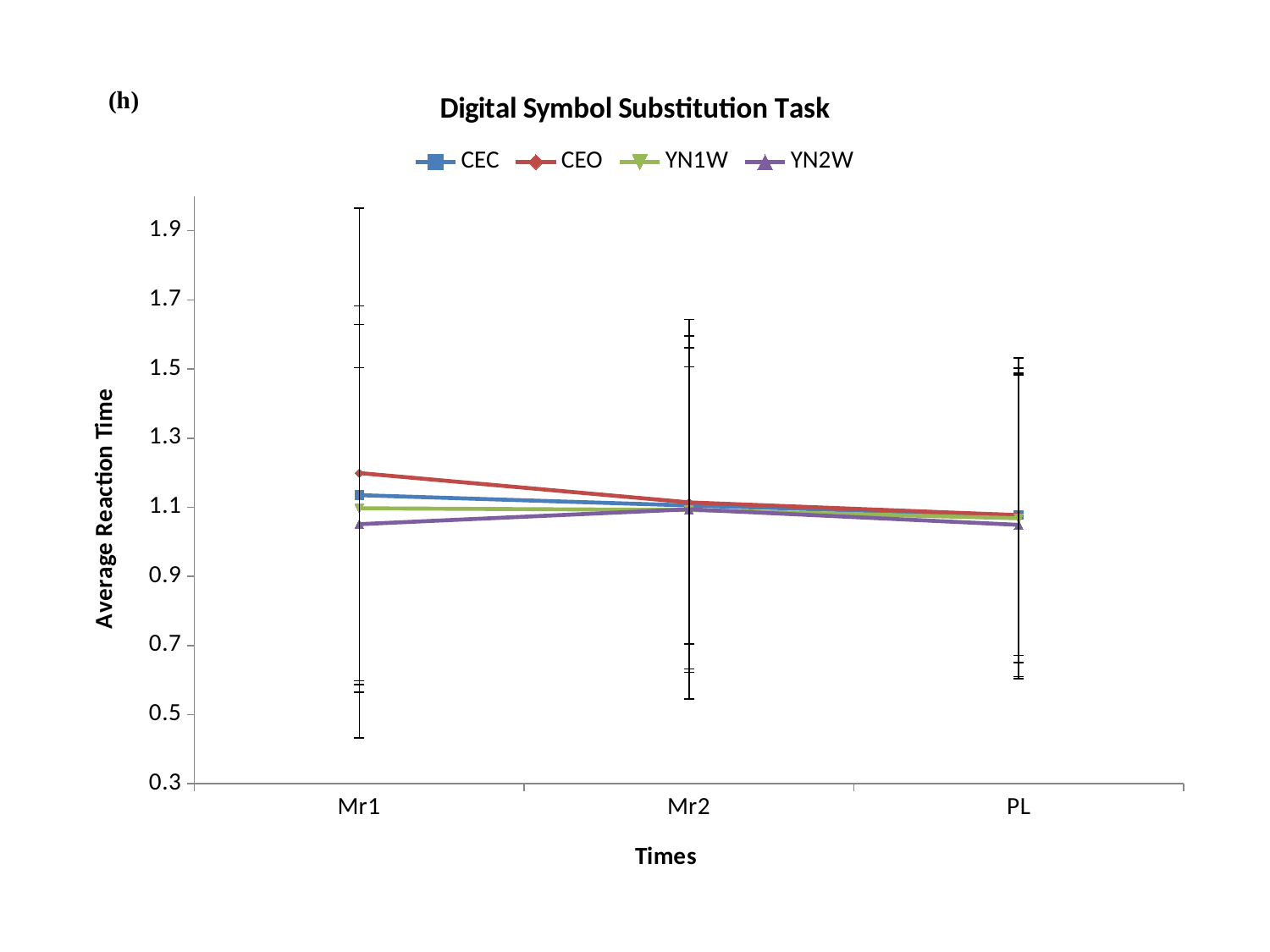

#### Chart: Digital Symbol Substitution Task
| Category | CEC | CEO | YN1W | YN2W |
|---|---|---|---|---|
| Mr1 | 1.135 | 1.199 | 1.097 | 1.051 |
| Mr2 | 1.105 | 1.114 | 1.092 | 1.094 |
| PL | 1.077 | 1.077 | 1.068 | 1.049 |

### Slide 9
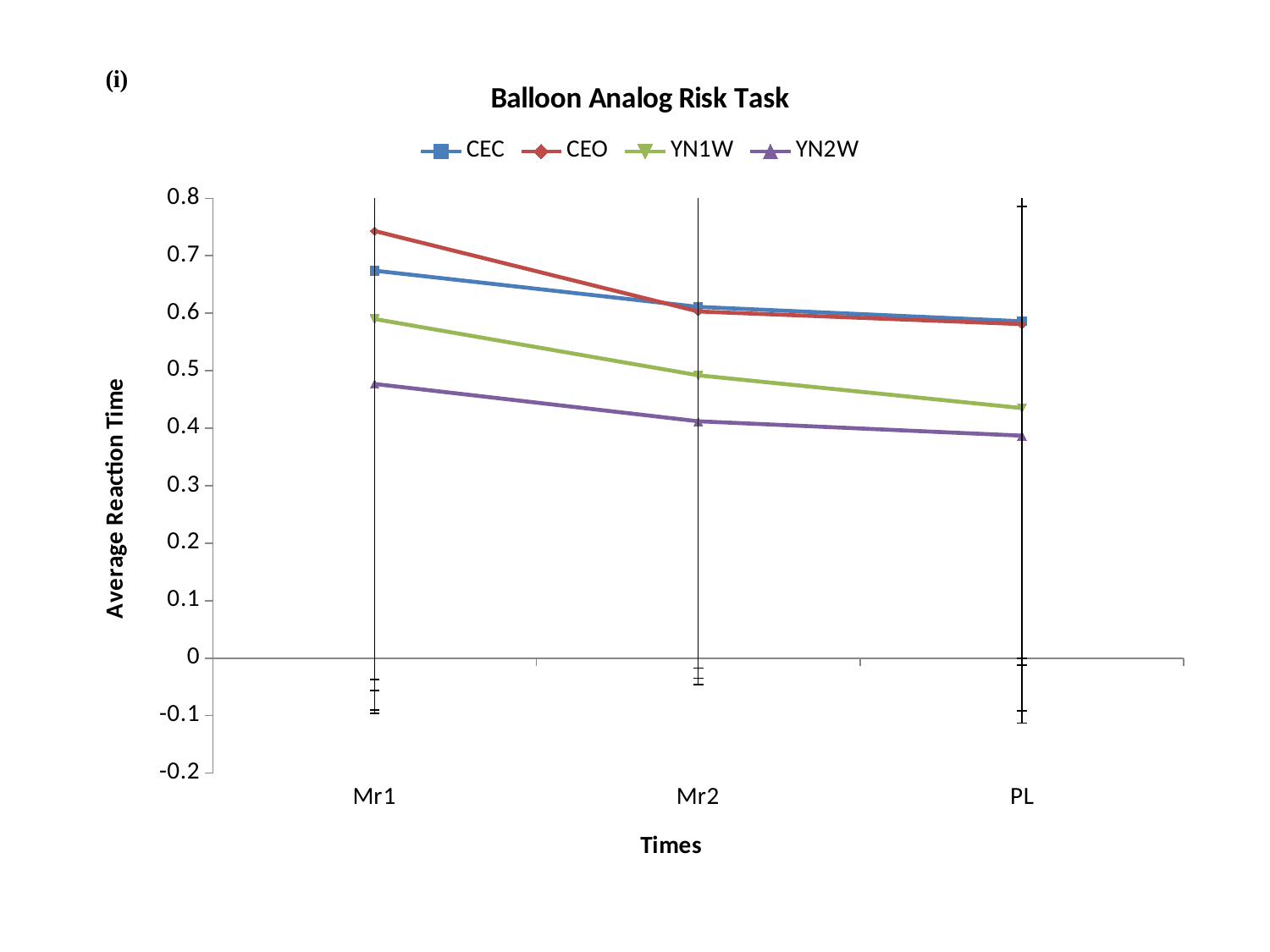

#### Chart: Balloon Analog Risk Task
| Category | CEC | CEO | YN1W | YN2W |
|---|---|---|---|---|
| Mr1 | 0.674 | 0.743 | 0.59 | 0.477 |
| Mr2 | 0.611 | 0.603 | 0.492 | 0.412 |
| PL | 0.586 | 0.581 | 0.435 | 0.387 |

### Slide 10
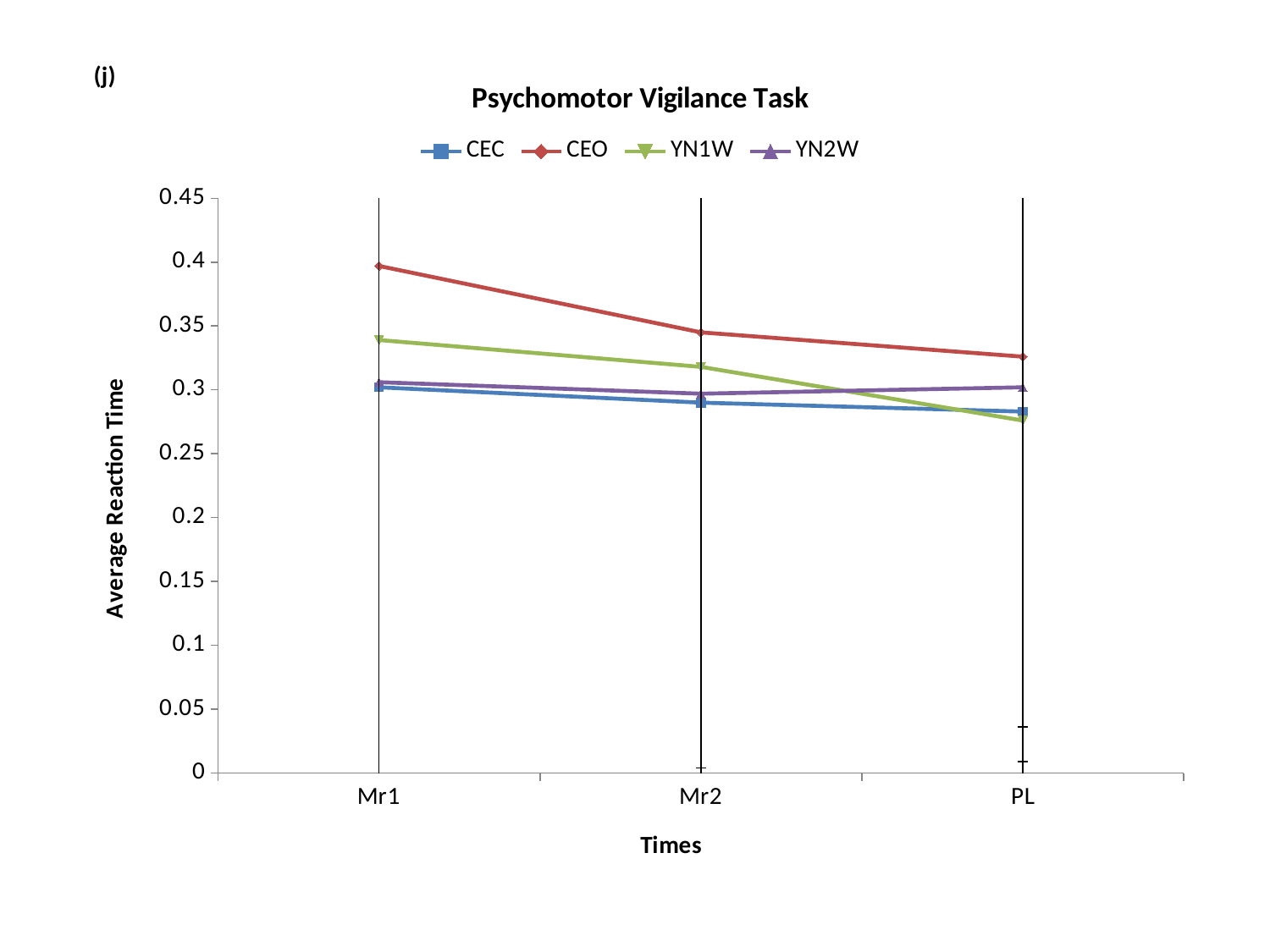

#### Chart: Psychomotor Vigilance Task
| Category | CEC | CEO | YN1W | YN2W |
|---|---|---|---|---|
| Mr1 | 0.302 | 0.397 | 0.339 | 0.306 |
| Mr2 | 0.29 | 0.345 | 0.318 | 0.297 |
| PL | 0.283 | 0.326 | 0.276 | 0.302 |
